# Geographical aggregation of asynchronous wastewater-based surveillance programmes, with application to Canada

**DOI:** 10.64898/2026.10.26.26366498

**Authors:** Warsame Yusuf, David Champredon

## Abstract

Wastewater-based surveillance (WBS) programmes increasingly monitor pathogens across large geographic areas, but aggregating asynchronous, heterogeneous site-level data into broader-scale indicators remains a methodological challenge. We developed a Bayesian hierarchical framework to estimate continuous latent wastewater concentration at broader geographic scales from asynchronous site-level observations. The model accounts for differences in measurement precision, catchment population size, and changes in site participation, while preserving temporal resolution and supporting real-time analysis. The framework was validated using simulated data and applied to wastewater concentration measurements of respiratory viruses collected between January 2024 and December 2025 from 25 sites in six provinces participating in Canada’s National Wastewater Monitoring of Pathogens programme. Application of the model to observational data generated continuous provincial and national concentration estimates. At the national level, latent wastewater concentrations showed moderate to strong agreement with corresponding clinical test positivity data (*R*^2^ = 0.65–0.93). Sensitivity analyses indicated that estimates were robust to shorter fitting windows and to the addition or removal of surveillance sites. This framework provides a statistically coherent and operationally practical approach for aggregating asynchronous wastewater observations into population-level surveillance indicators, with potential applications across pathogens and public health jurisdictions.

## 1 Introduction

During the COVID-19 pandemic, wastewater-based surveillance (WBS) became a popular tool for public health professionals worldwide to monitor the spread and prevalence of SARS-CoV-2 in jurisdictions, in which samples collected from treatment plants are tested (often with quantitative polymerase chain reaction (qPCR)) to measure the concentration of a pathogen in wastewater as a proxy of disease transmission in a community. Today, WBS is still used to monitor the spread of infectious pathogens that are shed in stool and/or urine of infected individuals. While the aim of WBS is to support public health surveillance, which can cover large geographic areas, the observed sample data are inherently localized and asynchronous. Aggregating those localized epidemiological signals into public health intelligence at a larger spatial scale is challenging. This same challenge arises across several scientific fields where aggregating localized, asynchronous observations to infer trends at higher geographic levels. One approach to this challenge is the use of Bayesian hierarchical models. Examples of this implementation include air-quality monitoring [1, 2], ecological surveillance [3], and clinical disease surveillance [4, 5]. In these examples, asynchronously gathered localized data are used to infer latent processes at broader geographic levels.

Larger WBS programmes that examine trends at national or subnational levels require data from multiple sampling locations (typically wastewater treatment plants) to represent the population of interest. In recent years, several countries have established large-scale wastewater surveillance programmes. In Canada, the National Wastewater Monitoring of Pathogens (NWMP) programme collects data from 44 treatment sites as of July 2026, covering approximately 30% of the Canadian population [6]. In the United States, the National Wastewater Surveillance System had expanded to over 1500 treatment sites as of December 2022, covering nearly 155 million people (*∼*45% of the population) [7]. Comparable surveillance programmes have also been implemented internationally in Europe and Asia [8, 9]. Many countries also have provincial or other subnational programmes that cover multiple sites within their respective jurisdictions [10, 11]. The growing scale of wastewater surveillance worldwide indicates that aggregation across sites is a central methodological problem in WBS.

As treatment plants differ in catchment size and sampling frequency, aggregating their data into higher-level concentration estimates poses several challenges. One approach used to aggregate wastewater data is to take a population-weighted average of observed sample concentrations from multiple sites. In this approach, the catchment population of each treatment plant is used to generate an aggregated mean concentration of a wider jurisdiction [6, 12]. This approach assumes sampling is done with the same frequency and on the same dates across sites (as missing values would change the denominator of the weighted average). Weekly averaging of individual sites is a method often used prior to aggregation [6, 13]. However, if sites are sampled several times in a week, weekly averages can reduce the resolution of the data time-series. Another method is to use linear interpolation to estimate concentration on non-sampling dates [14, 15, 16] prior to generating population-weighted mean concentration. While interpolation aligns sampling without reducing resolution, it assumes equal variance across sites in that concentrations are equally precise regardless of catchment size. Additionally, linear interpolation is retrospective in nature, as it requires subsequent data points to estimate concentration values in a time-series. Consequently, it provides limited support during real-time analysis of data. Previous studies have suggested that wastewater signals from sites with smaller catchment populations tend to have greater temporal variability than sites with larger catchments and, thus, may provide less precise estimates of temporal trends [17, 18].

We developed a statistical framework to aggregate viral wastewater concentration data from multiple sites to obtain estimates of latent wastewater concentration for higher geographic levels that encompass the selected sites. Within this framework, a Bayesian hierarchical model draws information from individual sampling sites (typically wastewater treatment plants) to estimate latent trends at broader geographic levels, using catchment population to infer the contribution of each treatment plant to the latent estimates of viral concentration. The model also assigns each site its own precision, so that concentrations measured at plants with larger catchments contribute more strongly to the latent estimate. By modelling site-specific precision and estimating latent concentration directly, the framework preserves temporal resolution, supports real-time analysis, and allows aggregation at multiple administrative levels (*e.g.,* regional, provincial, or national), addressing the limitations of weekly averaging and linear interpolation. We validate this modelling framework by testing it on simulated observational data, and then apply it to observational data collected from the Canadian NWMP programme, estimating provincial and national latent concentrations. We also test the robustness of this framework with the use of data windows of varying sizes and the addition and removal of sites to mimic the site-level operational changes commonly seen in WBS programmes. By addressing the limitations of existing methods, this framework provides a common approach to aggregating localized, asynchronous observations in WBS to support public health surveillance.

## 2 Methods

### 2.1 Data

The wastewater data come from the Public Health Agency of Canada (PHAC) National Wastewater Monitoring of Pathogens (NWMP) programme. Methods on how wastewater data in this programme are collected, processed, and tested at PHAC can be found in [19]. We retrieved viral concentration data from wastewater treatment plants in six provinces (British Columbia, Alberta, Manitoba, Ontario, Quebec, and Nova Scotia), selected as each contained multiple sites within the NWMP programme. The viruses selected were Influenza A, Influenza B, RSV, and SARS-CoV-2. Since RSV is generally reported as a single virus in traditional clinical surveillance but measured as subtypes A and B within the NWMP programme, reported sample concentrations were summed. Data were collected for 2 years, from 1 January 2024 to 31 December 2025. Not all sites included in the model had data that spanned the entire window of the study. Supplementary Table S3 presents the start and end dates of sites included in the model.

To compare latent estimates of wastewater concentration against clinical surveillance data, we used respiratory virus test positivity data from PHAC’s Respiratory Virus Detection Surveillance System (RVDSS), a national programme that collects respiratory testing data (number of tests performed and positive tests) from participating laboratories across Canada [20]. Provincial weekly test positivity (the proportion of positive tests) was extracted for SARS-CoV-2, Influenza A, Influenza B, and RSV, and restricted to the same provinces and study window used in the model. A national positivity time-series was derived as a population-weighted average of provincial positivity, using the same provincial population weights applied to the latent concentration estimates. For each virus-location pair, mean daily latent concentration was paired with clinical test positivity on matching dates, overlaid as a time-series, and the association summarized by the coefficient of determination (*R*^2^) from a linear regression of log_10_ positivity on log_10_ latent concentration. To ensure zero values are captured in the log regression, a small value close to zero was added to positivity values.

### 2.2 Statistical model

We used a hierarchical model to estimate a provincial wastewater index using a Bayesian random walk framework. This model contains two levels:

- the provincial level (first level of aggregation), where daily *latent* provincial concentration is informed by estimates of the previous day.
- the site level, where *observed* site concentrations inform latent daily provincial concentrations.

National concentration (and any higher level of aggregation) was obtained as a population-weighted average of the latent provincial estimates, rather than as an additional latent level. This parsimonious approach was preferred because the provincial process already provides daily estimates.

The latent provincial concentration at time *t* is modelled by a random walk:

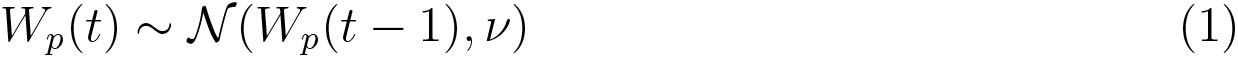

In this expression, latent provincial concentration estimates *W_p_*(*t*) are drawn by concentration estimates from the previous day *W_p_*(*t −* 1), with a standard deviation of *ν* that is shared across provinces.

The site level is based on a log-linear regression. For a given site *i* within a province *p* at time *t*, we define the log-concentration *W_p,i_*(*t*) = log(*w_p,i_*(*t*)) where *w_p,i_*(*t*) is the observed sample concentration. To model the relationship between site-level and provincial-level concentration, we assume that the mean of the log-concentration at the site level is linearly dependent from the provincial log-concentration:

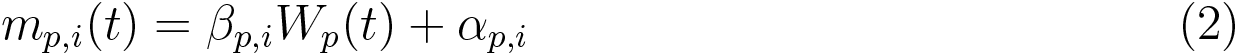

In this expression, *β_p,i_* represents the slope fraction in which mean site level concentration *m_p,i_* contributes to the provincial latent concentration *W_p_*, and *α_p,i_* represents site-specific random effects.

To account for variability and differences in catchment populations among the wastewater treatment sites, observed *W_p,i_*(*t*) was modelled using the mean concentration *m_p,i_*(*t*) obtained in Equation 2 using the following expression:

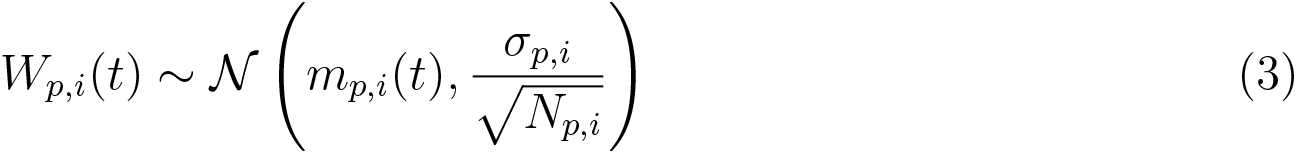

In this expression, observed site concentration *W_p,i_* is assumed to be normally distributed around the mean daily concentration *m_p,i_*. The standard deviation *σ_p,i_*is site-specific and is scaled by the square root of a site’s catchment population *N_p,i_*.

This allows each site its own measurement variability (σ^2^_*p,i*_), scaled by catchment population so that variance (σ^2^_*p,i*_/*N_p,i_*) decreases, and precision increases, with population size.

To aggregate latent concentration from several provinces, daily latent national concentration can be expressed as a population weighted average of the provincial estimates:

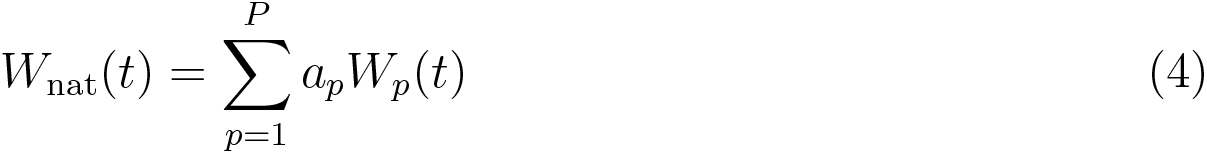

where *W_p_*represents the latent provincial concentration obtained in Equation 1, and *a_p_*represents the proportion of the total modelled population residing in province *p*. Because not all Canadian provinces or their full populations were covered, modelled provincial and national populations were defined as the summed catchment populations of included sites in each respective geographic level.

### 2.3 Model priors

Supplementary Table S1. summarizes the priors used in the hierarchical model. For province-specific and site-specific parameters, identical initial values were assigned across all sites and provinces.

### 2.4 Model implementation

The hierarchical model was implemented in R (version 4.4.3) using the nimble package (version 1.4.0) [21, 22]. To evaluate the validity of this model, simulated observation and latent provincial data were simulated using predefined values specified in Supplementary Table S2.

The model was fitted using a Markov chain Monte Carlo (MCMC) algorithm, iterated 500000 times across four chains, with 250000 iterations discarded as “burn-in”.

Estimates of the posterior distribution of the parameters used in the model were retrieved. To quantify between-site variability in the site-level parameters, we computed a coefficient of variation (CV) for each parameter within each virus. Because the site intercepts (*α*) are centred near zero (Supplementary Table S1), the conventional CV (standard deviation divided by the mean) was likely to produce unstable results. We therefore defined the CV as the standard deviation of the per-site posterior means divided by their mean absolute value, which remains well-defined when the signed mean approaches zero.

To confirm sufficient convergence of the MCMC algorithm chains, we calculated the potential scale reduction factor (PSRF) using the Gelman-Rubin statistic [23].

### 2.5 Ethics approval

Because this study relies exclusively on the secondary use of anonymous information, and the analysis does not generate identifiable information, it did not require a research ethics board review in accordance with Article 2.4 of the Tri-Council Policy Statement: Ethical Conduct for Research Involving Humans (TCPS 2) [24].

## 3 Results

### 3.1 Study population and observed data

Wastewater concentration data of SARS-CoV-2, Influenza A, Influenza B, and RSV were collected from 25 sites across six provinces, representing a total community population of approximately 12.2 million, or 32.8% of the Canadian population [25]. Supplementary Table S3 lists the start and end dates of each site included in the model.

Time-series plots of the observed wastewater concentration data are presented in Supplementary Figure S4.

### 3.2 Validation of hierarchical model

To validate the hierarchical model, a simulated dataset consisting of known latent concentrations at each level (site, provincial, national) and parameter estimates (*α*, *β*, *σ*, *ν*), was generated. Results of this validation are shown in Supplementary Figures S1-S3 and illustrate that the model successfully recovered latent concentration estimates and the parameter values defined in the simulation. National concentration estimates derived from the population-weighted aggregation of latent provincial estimates were also consistent with simulated values.

### 3.3 Estimates of model parameters

Posterior distributions of the parameters used in both the random walk expression and the log-linear likelihood expression can be found in Supplementary Figure S5. Supplementary Table S4. presents estimates of the coefficient of variation (CV) of the site-level parameters that revealed differing degrees of between-site concordance. Parameter *β* was highly consistent across sites for all four viruses (CV *∼*0.05–0.08), indicating a stable relationship between site-level and provincial concentration. Parameter *σ* showed moderate between-site variation (CV *∼*0.49–0.55), while *α* showed considerable variation (CV *∼*1.16–1.28), suggesting site-specific differences in baseline concentration. Estimates of CV for each parameter were relatively consistent between viruses, indicating between-site differences were structural rather than virus-specific.

When examining estimates of parameter *ν* (the standard deviation shared across provinces) in Supplementary Figure S5, SARS-CoV-2 showed the smallest point estimate and narrowest credible intervals while both Influenza A and Influenza B showed the highest point estimates and widest credible intervals.

### 3.4 Hierarchical model diagnostics

The results in Supplementary Figures S6 and S7 show that all parameters used in the model had a potential scale reduction factor (PSRF) below 1.1, and most parameters below 1.02, indicating that the four chains had satisfactorily converged.

### 3.5 Latent estimates of provincial and national concentrations

Estimates of latent provincial wastewater concentration derived from the model are shown in Figure 1, and latent national wastewater concentration, derived by population-weighted averaging of the provincial posteriors, is shown in Figure 2. In Figure 3, we present each province’s latent trajectory with the national average.

**Figure 1:**
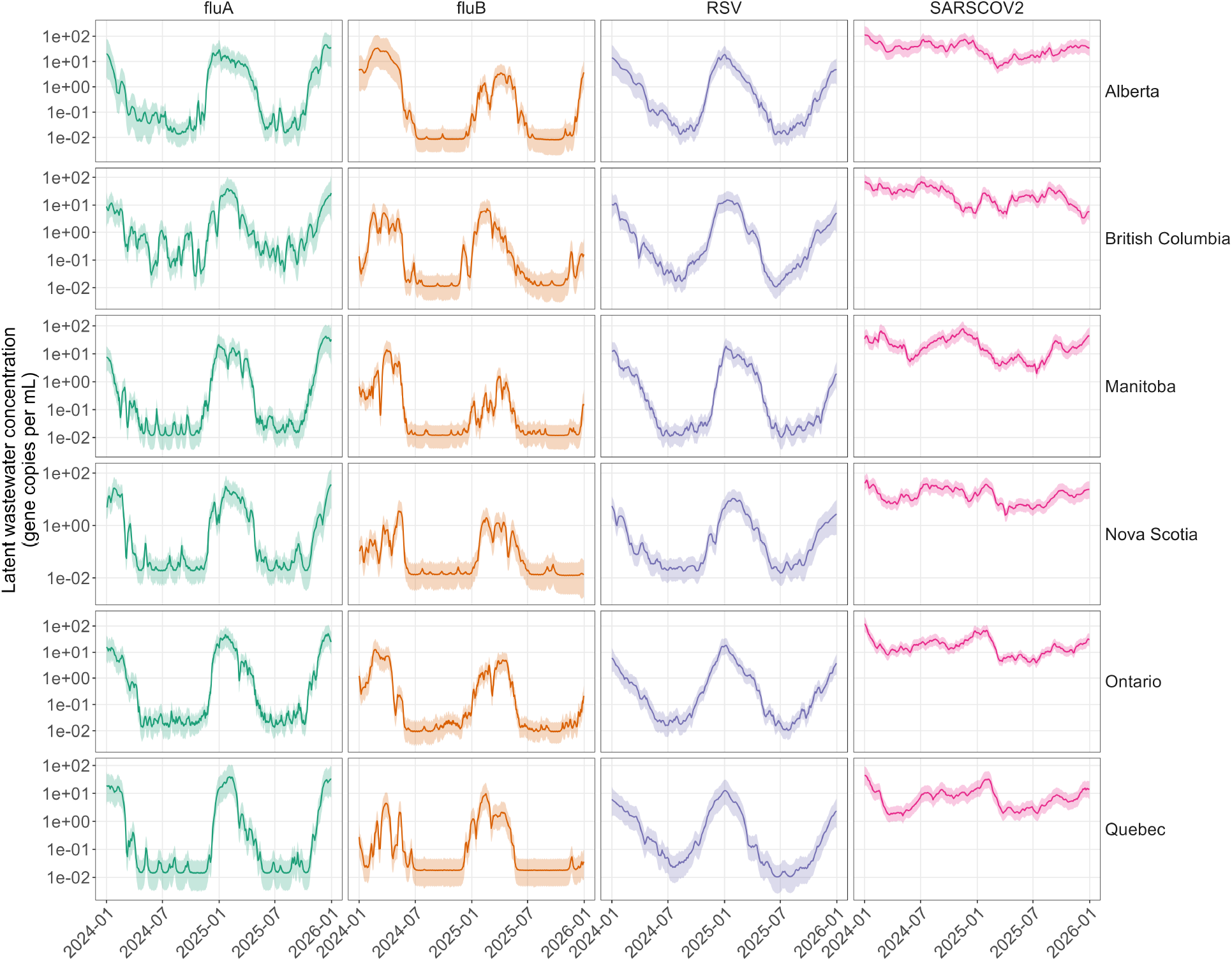
Estimates of latent provincial wastewater concentration for SARS-CoV-2, Influenza A, Influenza B, and RSV. Each subplot represents the time series of viral concentration for each virus in the six provinces, along with its 95% credible interval. The horizontal axis represents the latent sampling date, while the vertical axis represents the latent concentration on the log-scale.

**Figure 2:**
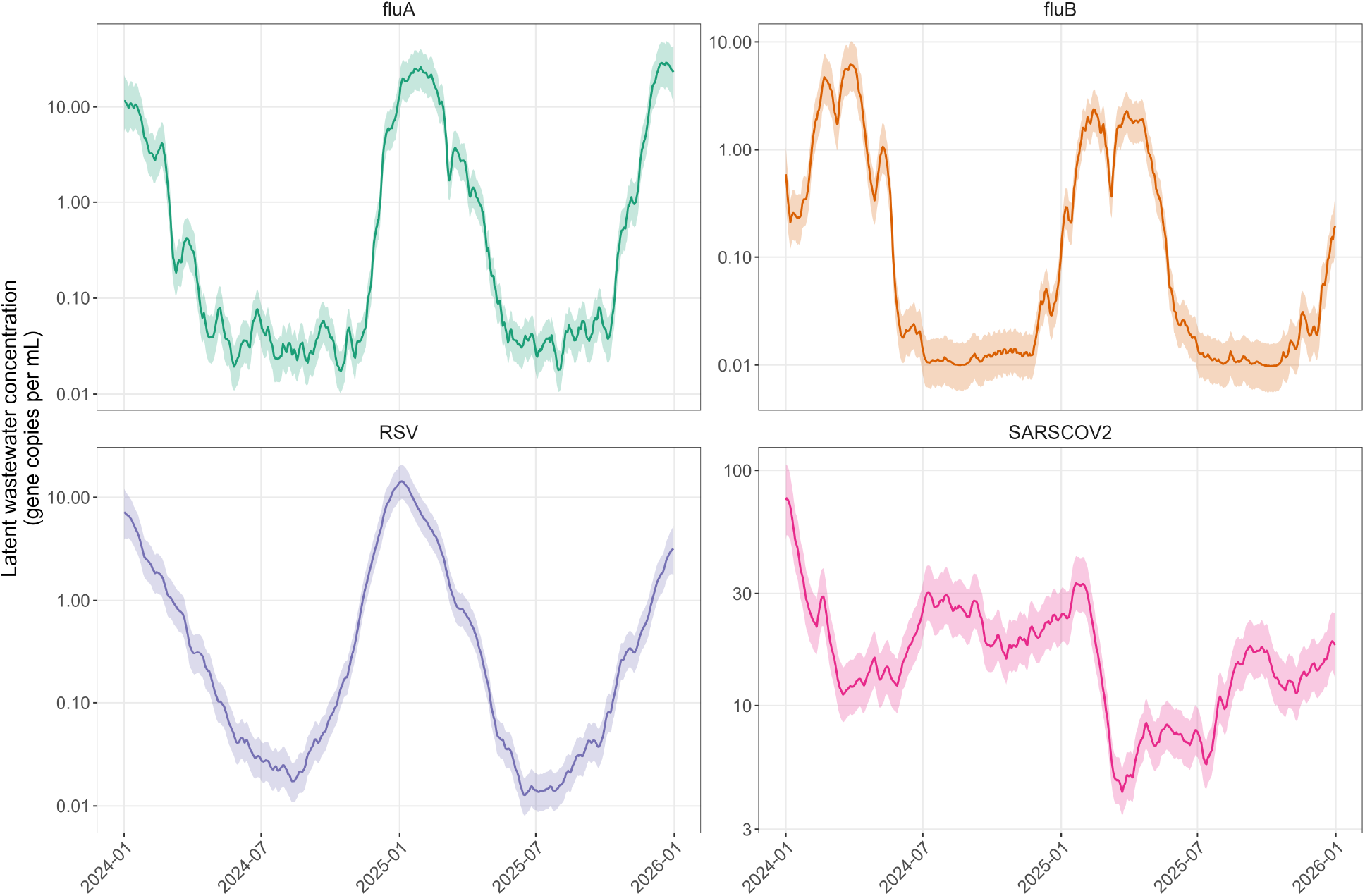
Estimates of latent national wastewater concentration for SARS-CoV-2, Influenza A, Influenza B, and RSV, derived from the posterior estimates of latent provincial concentration. Each subplot represents the time series of latent viral concentration for each virus, along with its 95% credible interval. The horizontal axis represents the latent sampling date, while the vertical axis represents the latent concentration on the log-scale.

**Figure 3:**
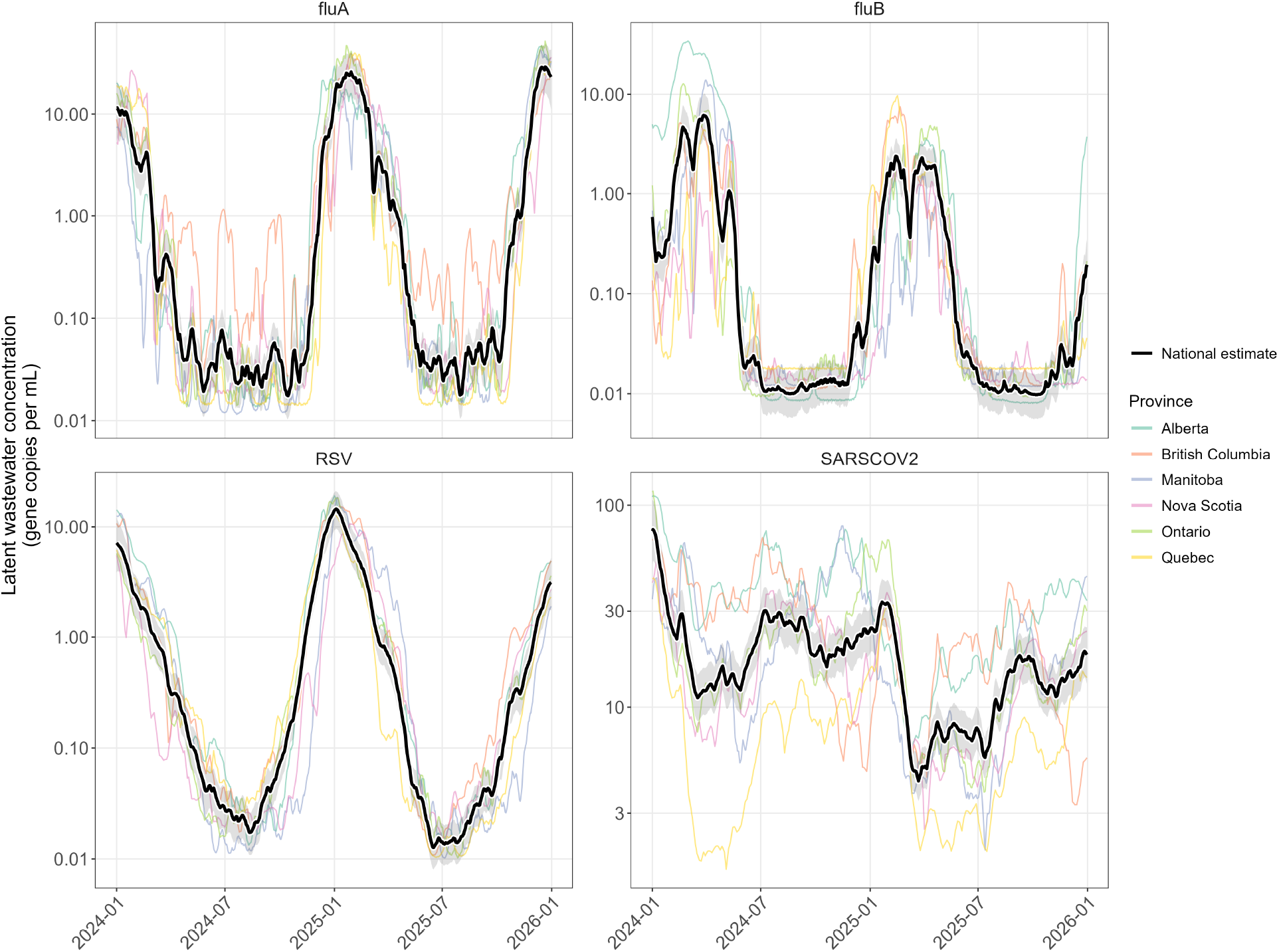
Estimates of latent national wastewater concentration for SARS-CoV-2, Influenza A, Influenza B, and RSV, with mean latent concentration for each province. Each subplot represents the time series of viral concentration for each virus. Black curve represents the national latent concentration with its 95% credible interval shaded in grey. The various coloured curves represent the mean latent concentration derived from the hierarchical model. The horizontal axis represents the latent sampling date, while the vertical axis represents the latent concentration on the log-scale.

### 3.6 Comparison to clinical surveillance data

Figure 4 and Supplementary Figure S8 present time-series plots that overlay latent wastewater concentration with clinical test positivity at the national and provincial level for SARS-CoV-2, Influenza A, Influenza B, and RSV. While the two data sources are on different scales (particularly for SARS-CoV-2), both figures present similar shapes, suggesting good temporal alignment across each virus-location pair.

**Figure 4:**
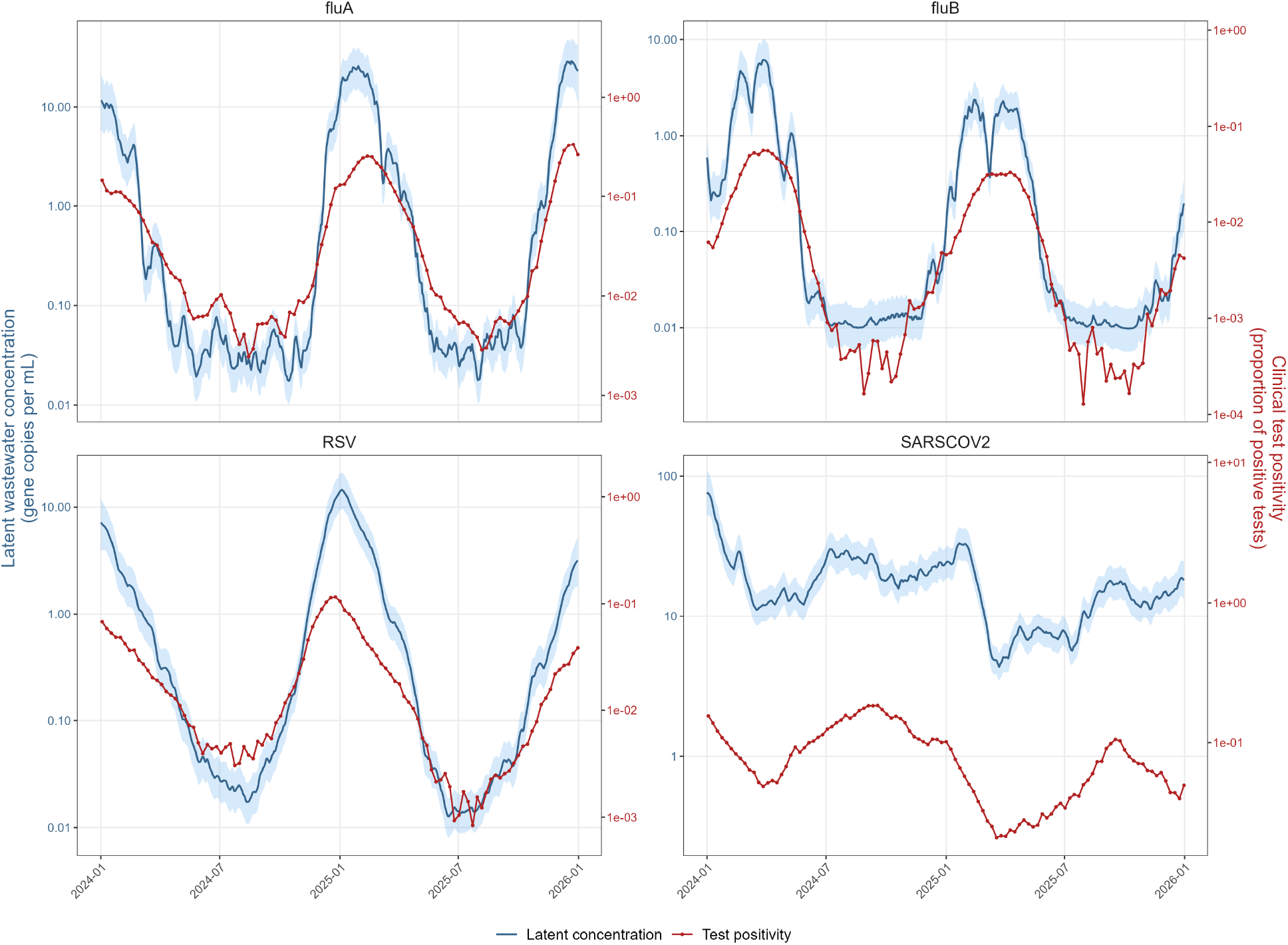
Comparison of latent national wastewater concentration and clinical test positivity for SARS-CoV-2, Influenza A, Influenza B, and RSV. Each subplot represents the time series for a virus, with latent concentration (blue, left axis) shown as the posterior mean and 95% credible interval, and clinical test positivity (red, right axis) shown on matching weekly dates. The two series are displayed on separate axes to emphasize that wastewater concentration and test positivity are measured on different scales.

To quantify the association, Supplementary Figures S9-S10 present scatter plots with estimates of *R*^2^ for each virus-location pair. The association was moderate to strong for Influenza A, Influenza B, and RSV (national *R*^2^ = 0.87–0.93, provincial: 0.44–0.92), and weaker for SARS-CoV-2 (national *R*^2^ = 0.65, provincial: 0.39–0.72).

### 3.7 Sensitivity analyses

#### 3.7.1 Comparison of 1-year vs 2-year window sizes

Using a subset of sites with full-window observations, latent provincial estimates from the full 2-year window were compared with estimates from two stitched 1-year windows (Supplementary Figure S11 and Table S5). Estimates were similar at any given time point.

#### 3.7.2 Addition and removal of sites

Using the same subset, latent provincial estimates with sites added or removed partway through the window were compared with the full-site estimates in Supplementary Figure S12 and Table S6. Estimates remained similar, with divergences localized to the periods following each event, and no event caused the model to fail or produce implausible estimates, demonstrating that the framework supports dynamic site addition/removal while maintaining a coherent provincial signal.

## 4 Discussion

Wastewater-based pathogen surveillance is inherently localized. Unlike conventional epidemiological indicators, such as case counts, its primary measurement unit (gene copies per litre) is not readily amenable to aggregation across larger geographic scales, including regional or national levels. Existing analytical approaches present important limitations that constrain the integration of this emerging surveillance framework into large-scale public health surveillance systems.

This study presents a statistical framework for aggregating wastewater measurements from multiple asynchronously sampled sites into coherent surveillance indicators at broader geographic scales. Unlike simple population-weighted averages or interpolation-based approaches, the framework simultaneously accounts for differences in sampling schedules, site-specific uncertainty, and catchment population size while generating continuous latent estimates of wastewater concentration. Although demonstrated using data from Canada’s National Wastewater Monitoring of Pathogens (NWMP) programme, the framework is applicable to wastewater surveillance systems operating at multiple administrative scales. Using data from the NWMP programme, we estimated latent concentrations of Influenza A, Influenza B, RSV, and SARS-CoV-2 across six Canadian provinces and subsequently generated national-level concentration estimates through population-weighted aggregation of provincial latent estimates.

Our statistical framework uses a random walk expression to infer latent provincial concentration in a time-series from observed concentration retrieved from wastewater treatment plants within each province. A random walk reflects the expectation that latent wastewater concentration varies smoothly rather than abruptly between consecutive days. This is consistent with the fact that measured concentrations represent aggregate faecal shedding from infected individuals, which can persist over several days and thereby smooth daily fluctuations in the signal [26].

In this context, latent concentration should not be interpreted as a directly observed measurement or an estimate of disease incidence. Rather, it represents a statistical reconstruction of the underlying wastewater signal after accounting for measurement noise, irregular sampling schedules, and differences between monitoring sites. This latent process provides a standardized surveillance indicator that can be compared across geographic levels and updated as new observations become available.

Catchment population was incorporated as a determinant of measurement precision (Equation 3), with uncertainty decreasing as population increases. Previous studies have shown that smaller catchments produce more variable signals while larger catchments yield more stable aggregates and stronger associations with epidemiological metrics [17, 27]. Because the random walk provides latent daily concentration estimates at the provincial level, national concentrations were derived by population-weighted averaging rather than including a national latent level in the hierarchical model. Preliminary analyses showed an additional national latent layer increased complexity and uncertainty without changing national trends.

Our simulation validation demonstrated that the hierarchical model successfully recovered latent provincial and national concentration signals from simulated site-level data. Simulated values of provincial and national concentration fell within the credible intervals of the recovered estimates, while the true parameter values (*α*, *β*, *ν*) were successfully recovered by the posterior distributions. These results provide evidence that the framework is capable of reconstructing latent concentration trajectories from asynchronous surveillance observations.

Application of the model to the NWMP dataset produced continuous provincial and national concentration estimates for all four viruses despite differences in sampling schedules and site participation over time. For seasonal viruses (Influenza A, Influenza B, RSV), credible intervals widened during periods of minimal wastewater activity, where measurements near assay detection limits are less informative. Comparison of provincial and national estimates illustrates the value of geographically aggregated surveillance indicators. Influenza A, Influenza B, and RSV displayed relatively concordant provincial trajectories, suggesting substantial synchronization of seasonal respiratory virus activity across jurisdictions. In contrast, SARS-CoV-2 exhibited greater heterogeneity among provinces, highlighting how aggregation may obscure localized epidemiological dynamics when transmission patterns differ across regions.

Our comparison of latent concentration estimates with clinical surveillance data provided several key findings that supported the model’s validity. Despite being on different scales, latent concentration tracked clinical test positivity well across the four viruses, in both in-season and inter-season periods. During in-season periods, latent wastewater concentration increased at a higher rate than clinical test positivity. This may reflect that wastewater captures viral material from infected individuals connected to sewer systems, including those with mild or asymptomatic infections who are less likely to be tested, whereas test positivity is additionally constrained by testing behaviour and capacity. During inter-season periods, where several province-virus pairs presented zero values for test positivity, latent wastewater concentration provided non-zero estimates, highlighting possible limitations of clinical testing outside of typical respiratory seasons, where latent estimates of concentration can serve as a proxy for clinical data. The quantified associations were moderate to strong for Influenza A, Influenza B, and RSV, and weaker for SARS-CoV-2, consistent with its greater heterogeneity. As the primary objective of this study was to present the framework for aggregating wastewater data, a detailed analysis of the wastewater-clinical relationship was not pursued here. Future studies focused on fitting wastewater data to clinical data could use our hierarchical model to generate latent concentration estimates at higher geographic levels as inputs to such analyses. Nevertheless, this comparison indicates that the statistical framework has strong utility in supporting public health surveillance.

Compared with commonly used aggregation approaches, this framework offers several advantages. Weekly aggregation methods address asynchronous sampling but reduce temporal resolution. Interpolation-based approaches preserve resolution but require future observations and are therefore less suitable for real-time surveillance applications. In contrast, the hierarchical model can generate updated latent concentration estimates immediately upon receipt of new wastewater observations while preserving daily temporal resolution. This capability is particularly relevant for public health surveillance programmes that require timely situational awareness during evolving outbreaks.

We used 2 years of data to capture both in-season and inter-season activity of seasonal respiratory viruses. Sensitivity analyses demonstrated that shorter data windows produced similar latent concentration estimates, suggesting that operational implementations may use shorter windows to reduce computational burden while retaining comparable results. The hierarchical model can also accommodate the addition and removal of surveillance sites over time. This characteristic reflects the realities of operational wastewater surveillance, where sites may enter or leave surveillance networks because of logistical, financial, or laboratory considerations. The ability to incorporate dynamic changes in network composition without restructuring the model enhances its applicability to long-term surveillance programmes.

While the framework provides a practical approach for aggregating wastewater concentration estimates, several limitations should be considered.

Provincial and national estimates were generated despite partial coverage of the Canadian population, reflecting the current participation of wastewater treatment plants within the NWMP programme. Although the selected sites represent major population centres that participate (or have participated) in the NWMP programme, unmonitored communities may experience epidemiological patterns that differ from those captured by the surveillance network. Consequently, the estimated provincial and national indicators should be interpreted as representative of the monitored population rather than the entirety of each jurisdiction. Because population-weighted aggregation can allow larger sites to dominate the combined signal, trends in smaller communities may be better captured through site-level analysis than aggregation.

Several measurement factors were simplified. Catchment population was used as a proxy for measurement precision, though variability may also arise from sewer network characteristics, industrial inputs, laboratory procedures, and environmental conditions such as precipitation, flow, and in-sewer degradation [28, 29]. Observations near or below assay detection limits were handled by substitution. Future work could incorporate additional site-specific predictors of uncertainty, standardized environmental metadata, and a more formal treatment of left-censored observations, though obtaining consistent metadata across sites may be challenging.

The model assumes a common temporal variance parameter (*ν*) across provinces, which improved model parsimony and computational efficiency, but may not reflect jurisdiction-specific dynamics. The model also does not explicitly address the modifiable areal unit problem (MAUP), where the definition of spatial boundaries can influence aggregated estimates [30, 31]. Future work could examine province-specific temporal variance structures or alternative formulations that allow temporal smoothness to vary geographically. Incorporating spatial correlation structures could potentially improve estimation, particularly in regions with sparse surveillance coverage.

Despite these limitations, the framework provides a practical, statistically coherent approach for aggregating asynchronous wastewater observations into surveillance indicators at broader geographic levels. By accounting for differences in sampling schedules, catchment population size, and site-specific uncertainty, it generates continuous latent concentration estimates while preserving temporal resolution. Although illustrated using Canadian wastewater data of respiratory pathogens, the approach is broadly applicable to surveillance networks operating in diverse geographic and epidemiological settings, offering a tool to support public health surveillance.

## 5 CRediT authorship contribution statement

**Warsame Yusuf** - Conceptualization, Data curation, Formal analysis, Methodology, Validation, Visualization, Writing - original draft, Writing - review and editing. **David Champredon** - Conceptualization, Formal analysis, Methodology, Validation, Writing - review and editing.

## 6 Data availability

The wastewater data are publicly available on the Public Health Agency of Canada (PHAC)’s wastewater surveillance dashboard https://health-infobase.canada.ca/wastewater/. The clinical test positivity data are also publicly available on PHAC’s respiratory virus surveillance dashboard https://health-infobase.canada.ca/respiratory-virus-surveillance/. The computer code for the statistical model will be available at https://github.com/phac-modelling-hub/wbs-aggregation-canada upon publication.

## Supporting information

Supplementary Material: Geographical aggregation of asynchronous wastewater-based surveillance programmes, with application to Canada

