## Supplementary Material: Geographical aggregation of asynchronous wastewater-based surveillance programmes, with application to Canada for "Geographical aggregation of asynchronous wastewater-based surveillance programmes, with application to Canada"

#### S1 Model priors

Table S1 presents the priors defined in the hierarchical model.

Table S1: Model priors

| Variable | Description | Distribution | Values |
|---|---|---|---|
| $\alpha_{p,i}$ | intercept of the log-linear regression | normal | $\mu = 0$, $\sigma = 0.3$ |
| $\beta_{p,i}$ | slope of the log-linear regression | normal | $\mu = 1$, $\sigma = 0.1$ |
| $\sigma_{p,i}$ | site-specific measurement error std. dev. | exponential | $\lambda = 0.25$ |
| $\nu$ | standard deviation shared across provinces | exponential | $\lambda = 0.2$ |
| $W_{p,1}$ | initial provincial concentration | normal | $\mu = \log(10)$, $\sigma = 1$ |

Table S2: Input parameters used to generate the simulated hierarchical dataset.

| Parameter | Symbol | Value |
|---|---|---|
| Number of provinces | $P$ | 6 |
| Time points | $T$ | 150 |
| Sites per province | $S$ | 5 |
| Total sites | $P \times S$ | 30 |
| Baseline concentration | $W_0$ | 10 |
| Site observation variability | $\sigma$ | 0.25 |
| Provincial deviation variability | $\nu$ | 0.15 |

### S2 Validation of hierarchical model

A simulated dataset was generated with predefined input parameters (Table S2). These inputs were used to simulate latent provincial and national concentration, site-level parameters ($\alpha$, $\beta$, $\sigma$, $\nu$), and site-level observation data.

Figure S1 presents the simulated site-level observation data used to fit the model.

Figure S2 overlays the model's recovered latent concentration estimates (posterior mean and 95% credible interval) against the simulated "truth" signal at both the provincial and national level, demonstrating that the model successfully recovered the latent concentration. Figure S3 presents the posterior distributions of the model parameters with the simulated truth values overlaid, confirming that the defined parameter values fell within the recovered posterior distributions.

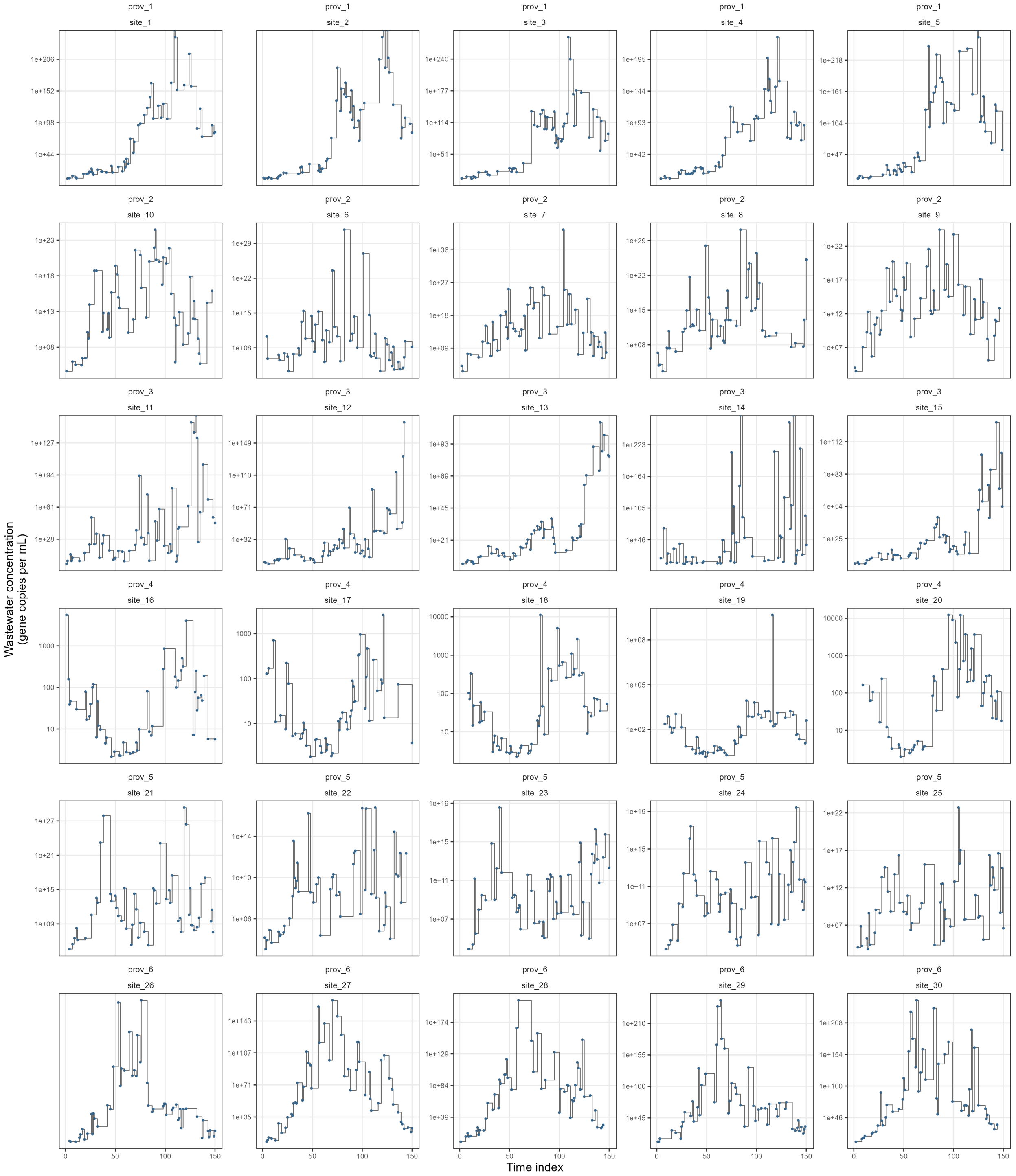


Figure S1: Simulated wastewater concentration data at each site used to fit the hierarchical model. Each subplot represents a time-series of simulated viral concentration for a site, arranged by province (rows) and site (columns). The x-axis represents the time index, while the y-axis represents the simulated concentration on the log-scale.

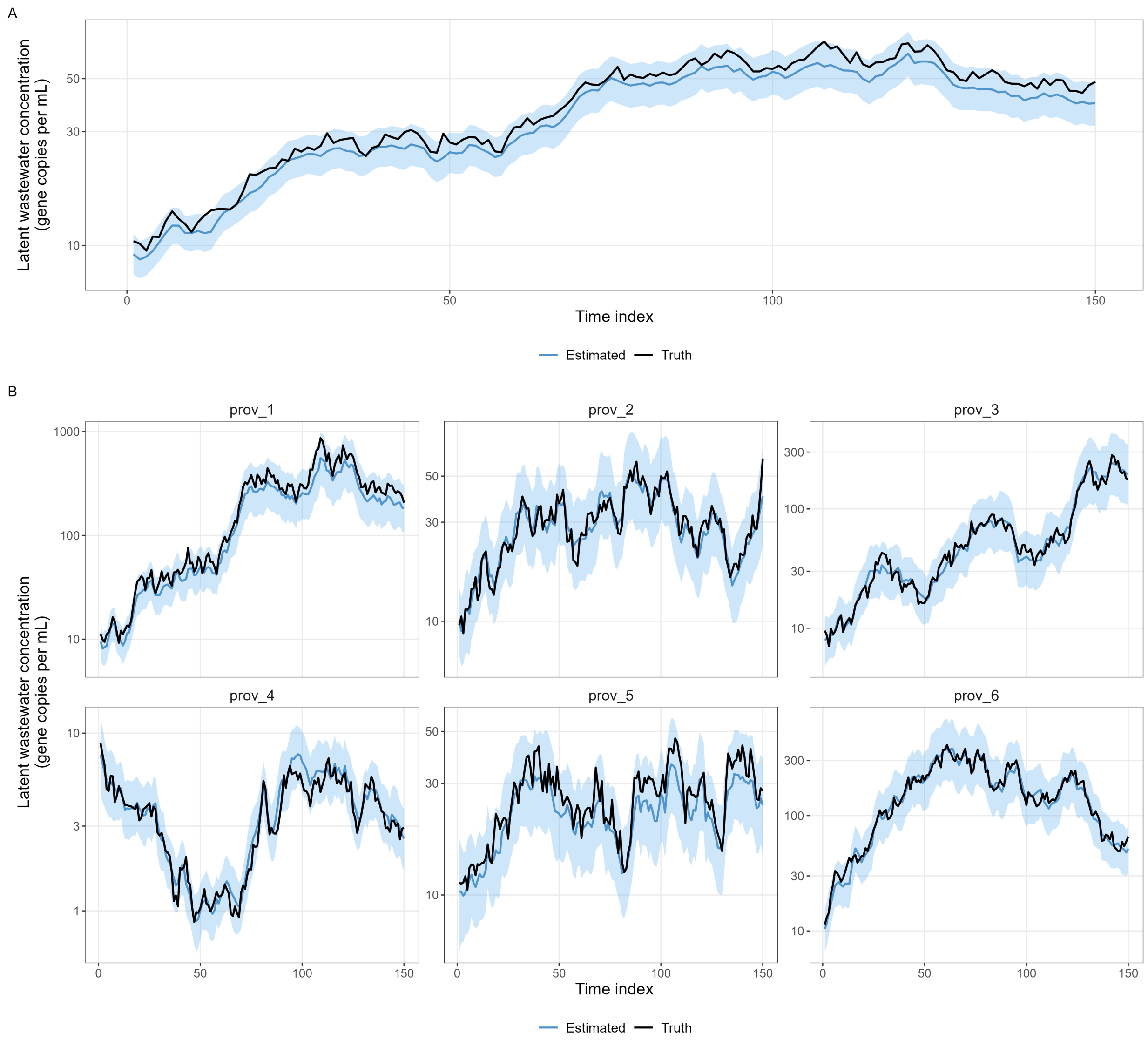


Figure S2: Recovered latent wastewater concentration (posterior mean, solid line, with 95% credible interval, shaded ribbon) overlaid with the simulated truth (dashed line). A) National concentration. B) Provincial concentration, faceted by province. The x-axis represents the time index, while the y-axis represents concentration on the log-scale.

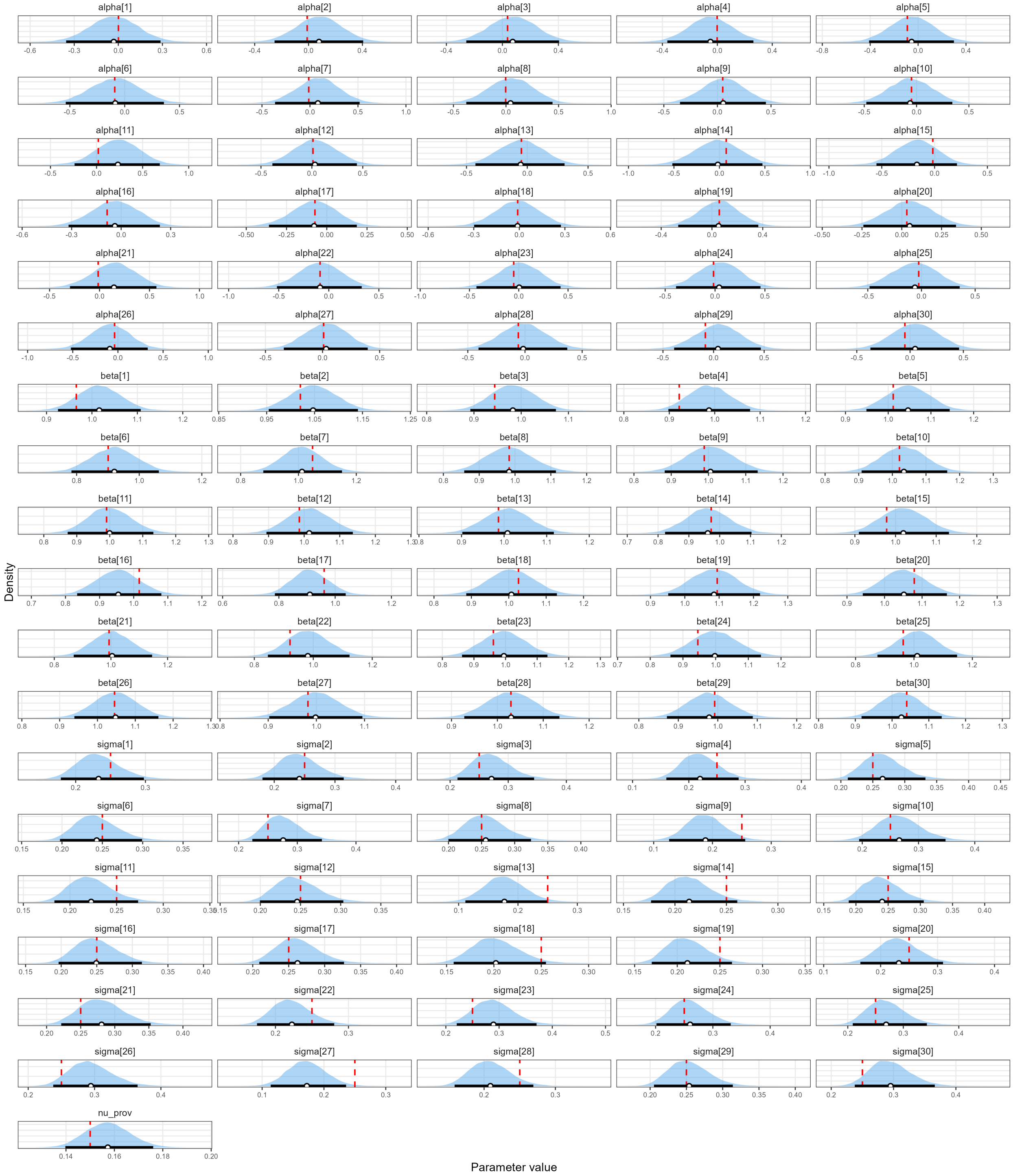


Figure S3: Posterior distributions of the model parameters ($\alpha$, $\beta$, $\sigma$, and $\nu$) recovered from the simulated dataset. Each subplot presents the posterior density (shaded), the posterior median and 95% credible interval (point and horizontal bar), and the simulated truth value (red dashed line). The x-axis represents the parameter value.

#### S3 Start and end dates of sites in model

Table S3 illustrates the first and last observation date for each wastewater site included in the hierarchical model. As mentioned in the methods, not all sites had observed data throughout the entire data window due to site inclusion and exclusion from the National Wastewater Monitoring of Pathogens programme. The number of observations by site vary due to sampling frequency (sites in the Winnipeg area are sampled five times per week, while others are sampled two times) and operational changes during the course of the data window.

Table S3: Wastewater site characteristics.

| Province | Site | Site ID | % prov. pop. | First obs. | Last obs. | N obs. |
|---|---|---|---|---|---|---|
| Alberta | Calgary Bonnybrook WWTP | CBB | 23.0 | 2024-06-05 | 2025-12-31 | 652 |
| Alberta | Calgary Fish Creek WWTP | CFC | 2.4 | 2024-06-05 | 2025-08-20 | 508 |
| Alberta | Calgary Pine Creek WWTP | CPC | 6.5 | 2024-06-05 | 2025-08-20 | 504 |
| Alberta | Edmonton Goldbar | EGB | 21.5 | 2024-01-02 | 2025-12-31 | 716 |
| British Columbia | Vancouver Annacis Island | VAI | 28.4 | 2024-01-04 | 2025-12-28 | 832 |
| British Columbia | Vancouver Iona Island | VII | 15.4 | 2024-01-04 | 2025-12-28 | 820 |
| British Columbia | Vancouver Lions Gate | VLG | 3.9 | 2024-01-04 | 2025-08-21 | 684 |
| British Columbia | Vancouver Lulu Island | VLI | 4.1 | 2024-01-04 | 2025-08-21 | 680 |
| British Columbia | Vancouver Northwest Langley | VNL | 0.9 | 2024-01-04 | 2025-08-21 | 684 |
| Manitoba | Winnipeg North End | WPN | 30.1 | 2024-01-01 | 2025-12-29 | 2068 |
| Manitoba | Winnipeg South End | WPS | 13.1 | 2024-01-01 | 2025-12-29 | 2048 |
| Manitoba | Winnipeg West End | WPW | 6.4 | 2024-01-01 | 2025-09-04 | 1700 |
| Nova Scotia | Halifax Dartmouth | HDA | 7.3 | 2024-01-01 | 2025-08-20 | 636 |
| Nova Scotia | Halifax Halifax | HHA | 15.4 | 2024-01-01 | 2025-12-31 | 832 |
| Nova Scotia | Halifax Millcove | HMC | 5.6 | 2024-01-01 | 2025-08-20 | 688 |
| Ontario | Kingston - Cataraqui Bay | KCB | 0.4 | 2024-10-18 | 2025-12-30 | 444 |
| Ontario | London-Admin | LON-A | 1.0 | 2024-10-05 | 2025-12-31 | 500 |
| Ontario | London-WIB | LON-W | 0.8 | 2024-10-05 | 2025-12-31 | 500 |
| Ontario | Peel G.E. Booth WWTP | PLG | 7.7 | 2024-09-23 | 2025-12-22 | 508 |
| Ontario | Toronto Ashbridges Bay | TAB | 10.2 | 2024-01-02 | 2025-12-30 | 836 |
| Ontario | Toronto Highland Creek | THC | 3.6 | 2024-01-02 | 2025-09-09 | 708 |
| Ontario | Toronto Humber | THU | 4.7 | 2024-01-02 | 2025-12-30 | 836 |
| Ontario | Toronto North Toronto | TNT | 1.4 | 2024-01-07 | 2025-09-09 | 687 |
| Quebec | Montreal North | MMN | 12.0 | 2024-01-03 | 2025-12-31 | 820 |
| Quebec | Montreal South | MMS | 12.2 | 2024-01-03 | 2025-12-31 | 828 |

### S4 Time series of observation data

Figure S4 illustrates the observed concentration data of the four viruses used in the hierarchical model.

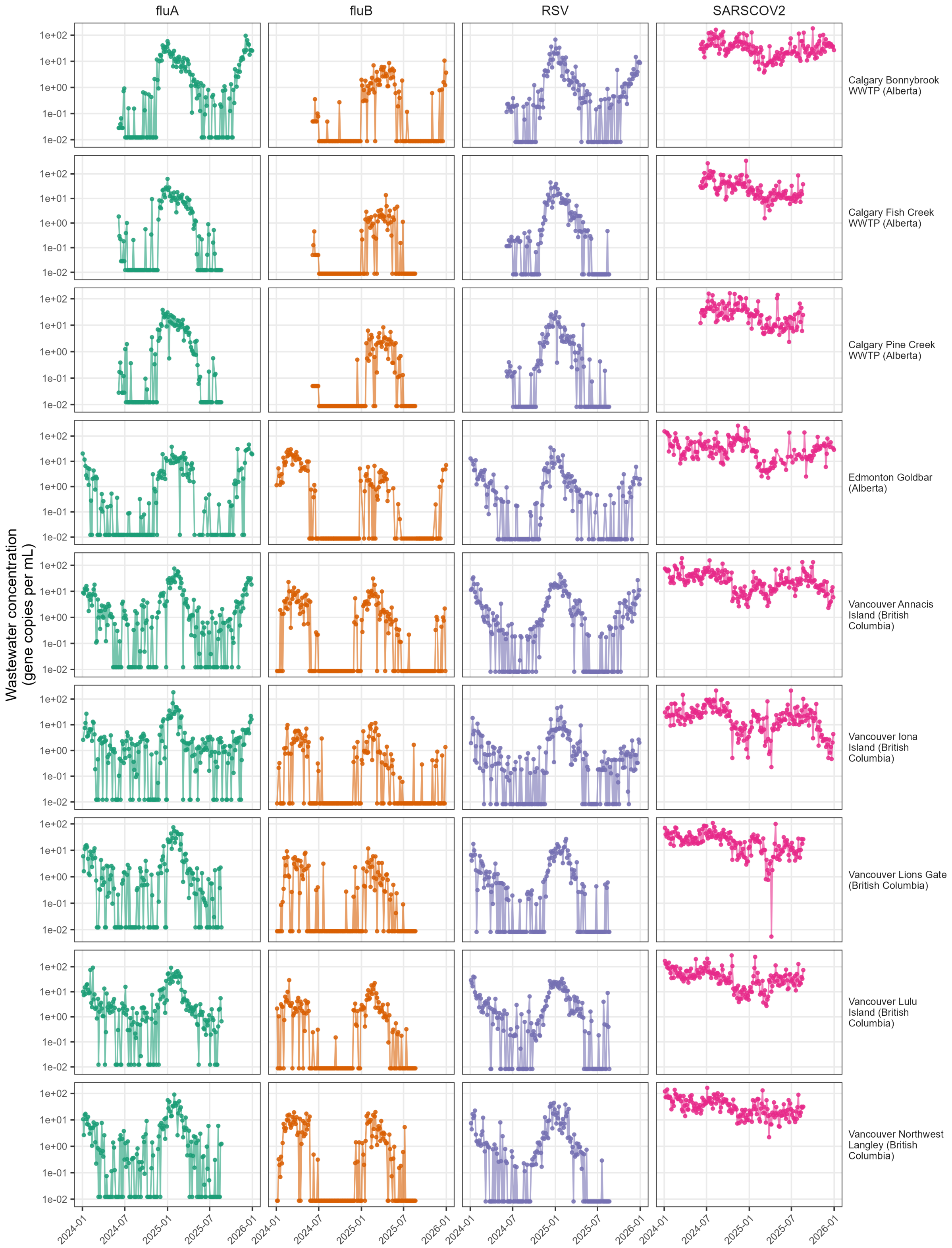


Figure S4: Observed wastewater concentration data of SARS-CoV-2, Influenza A, Influenza B, and RSV used in the model. Each subplot represents a time-series of viral concentration for each virus at each wastewater treatment plant. X-axis represents the date, while the y-axis represents the concentration on the log-scale.

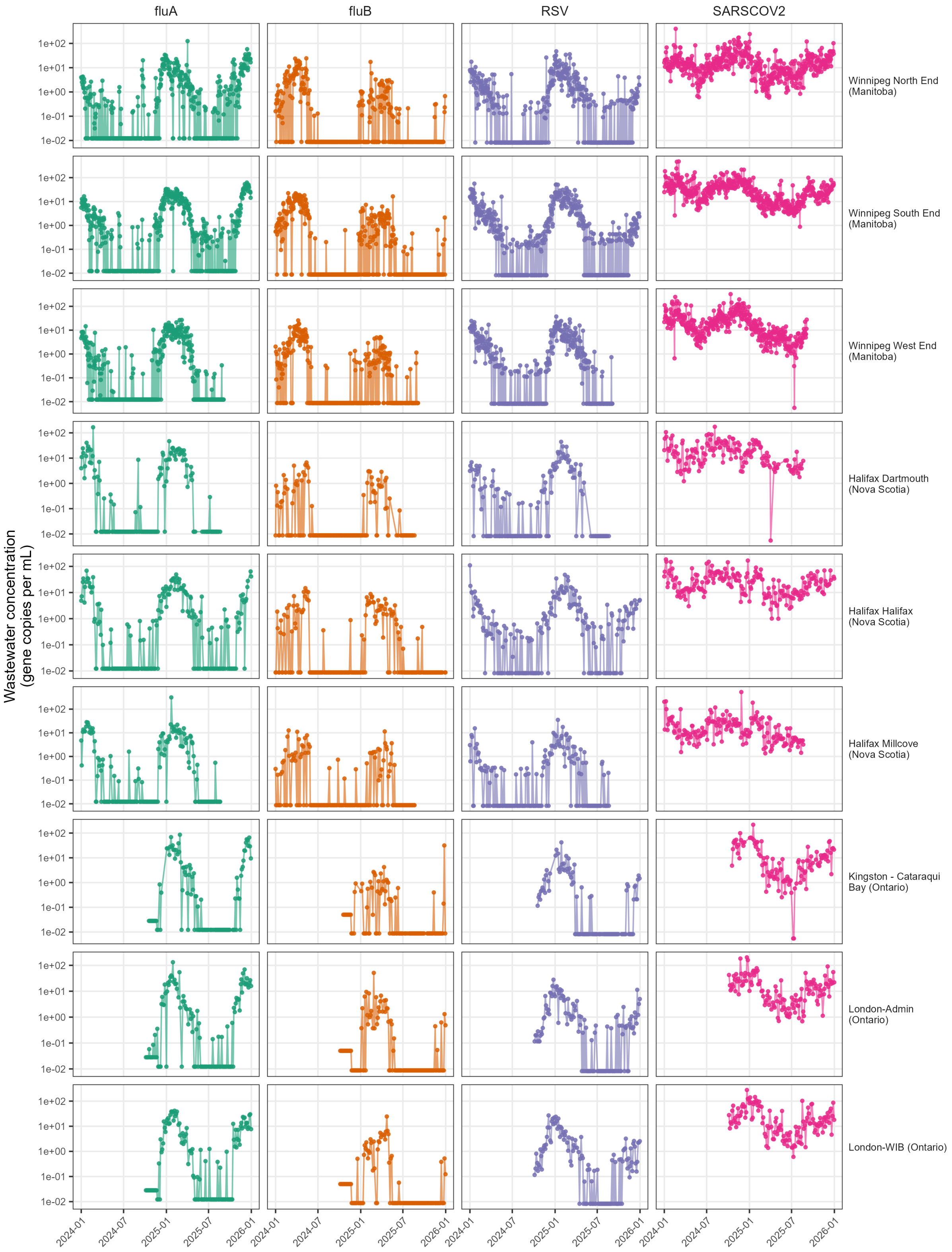


Figure S4: (continued)

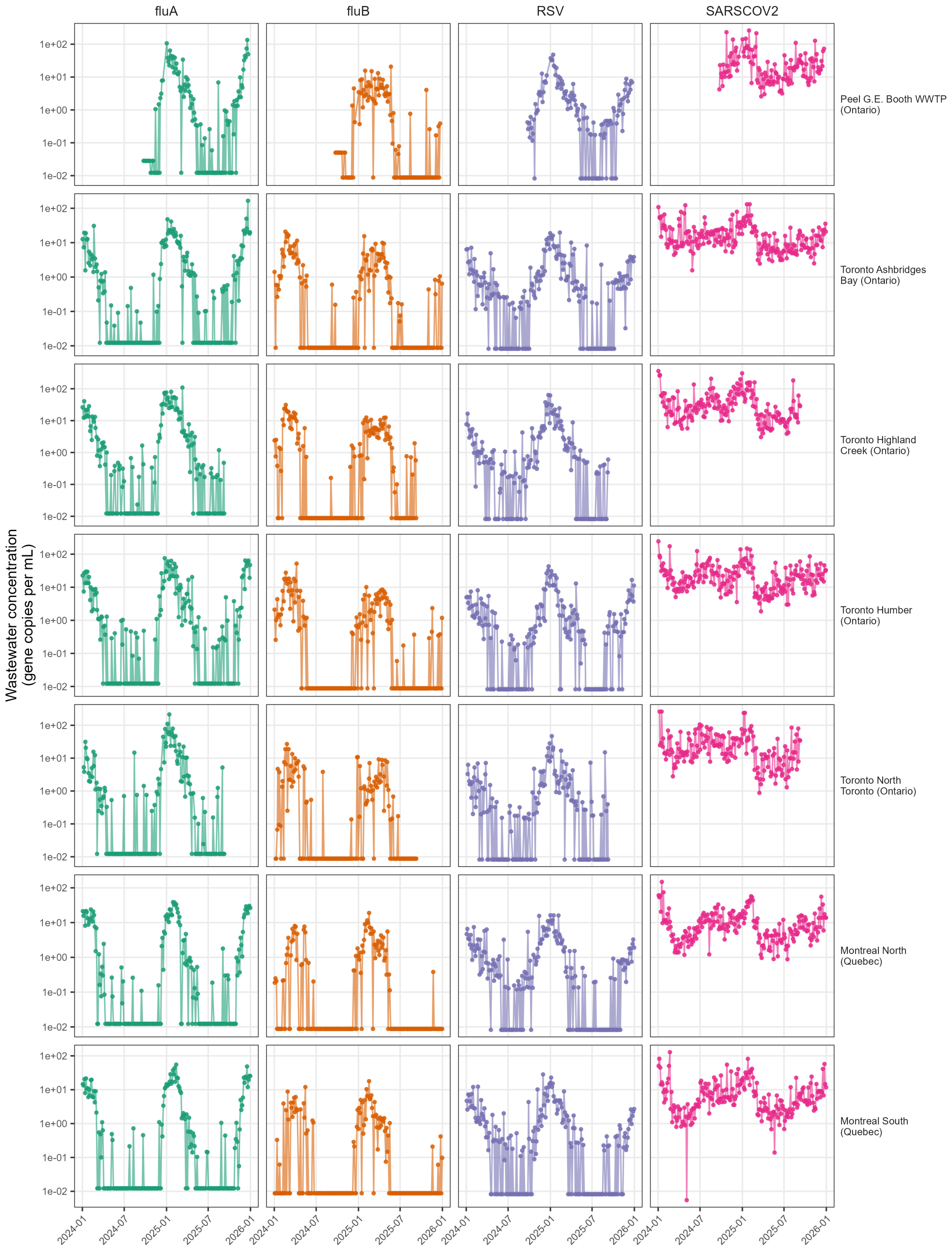


Figure S4: (continued)

Table S4: Coefficient of variation (CV) of site-level posterior parameter estimates ($\alpha$, $\beta$, $\sigma$) across sites, by virus. CV, coefficient of variation (standard deviation of per-site posterior means divided by their mean absolute value); SD, standard deviation; $|x|$, mean absolute value.

| Parameter | Virus | N sites | Mean $\|x\|$ | SD | CV |
|---|---|---|---|---|---|
| $\alpha$ | RSV | 25 | 0.310 | 0.388 | 1.252 |
| $\alpha$ | SARS-CoV-2 | 25 | 0.317 | 0.382 | 1.202 |
| $\alpha$ | Influenza A | 25 | 0.221 | 0.284 | 1.283 |
| $\alpha$ | Influenza B | 25 | 0.356 | 0.413 | 1.162 |
| $\beta$ | RSV | 25 | 1.004 | 0.053 | 0.053 |
| $\beta$ | SARS-CoV-2 | 25 | 1.007 | 0.077 | 0.077 |
| $\beta$ | Influenza A | 25 | 1.006 | 0.063 | 0.062 |
| $\beta$ | Influenza B | 25 | 1.008 | 0.080 | 0.079 |
| $\sigma$ | RSV | 25 | 1.253 | 0.646 | 0.515 |
| $\sigma$ | SARS-CoV-2 | 25 | 0.587 | 0.289 | 0.493 |
| $\sigma$ | Influenza A | 25 | 1.148 | 0.629 | 0.548 |
| $\sigma$ | Influenza B | 25 | 1.152 | 0.561 | 0.487 |

### S5 Posterior distributions of model parameters

Figure S5 illustrates the posterior distributions for the site-level parameters across the four viruses, as well as posterior estimates for the shared parameter $\nu$. Table S4 presents estimates of coefficient of variation (using mean absolute value) for the site-level parameters across the viruses.

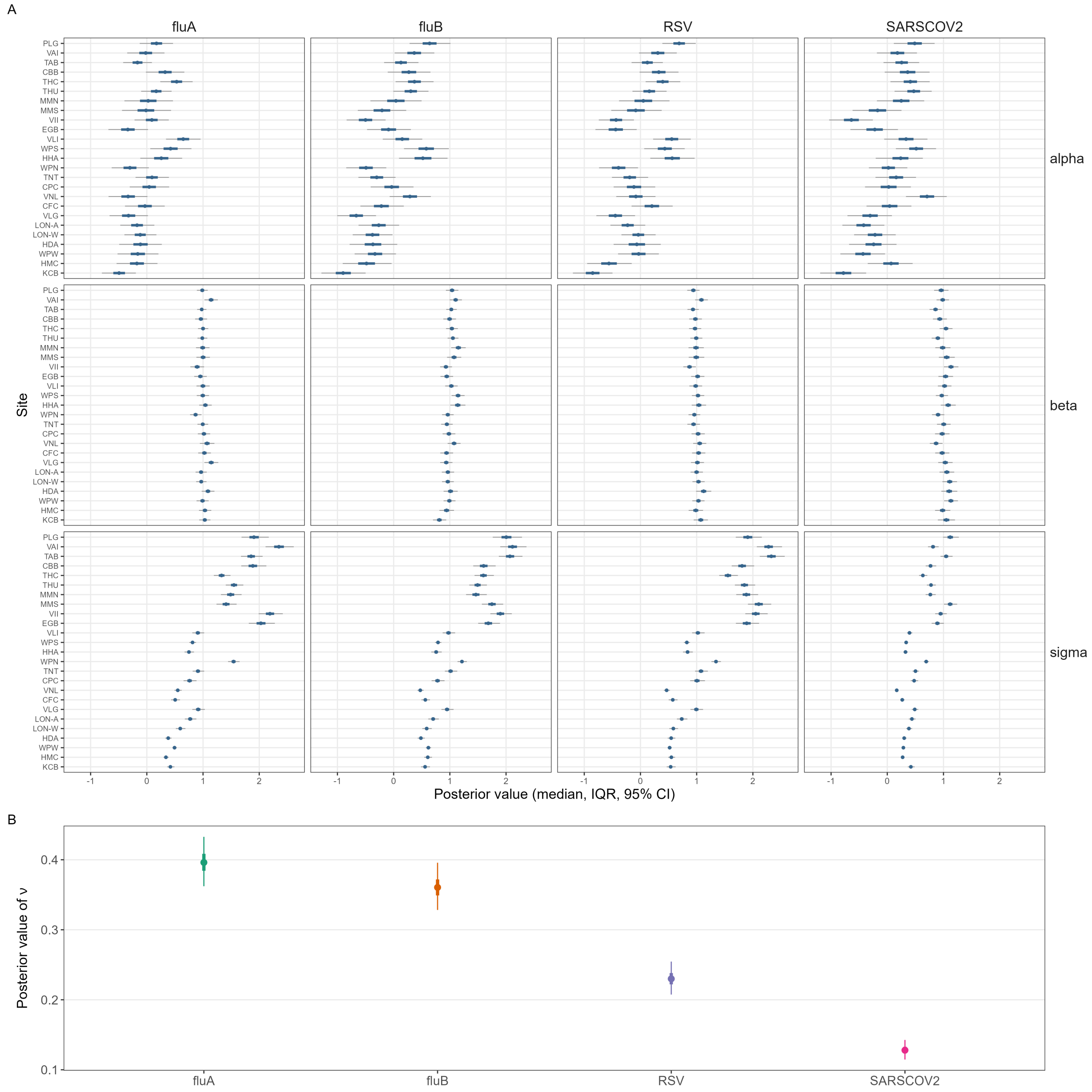


Figure S5: A) Posterior distribution for site-level parameters $\alpha$, $\beta$, and $\sigma$ for SARS-CoV-2, Influenza A, Influenza B, and RSV across all sites used in the model. Each subplot represents site estimates of median, 95% credible interval (CI), and interquartile range (IQR) for each virus and parameter. X-axis represents the posterior value, while the y-axis represents the site ID. B) Posterior distribution for $\nu$ for SARS-CoV-2, Influenza A, Influenza B, and RSV.

### S6 Gelman-Rubin statistic of convergences

Figure S6 illustrates the PSRF estimates for the site-level parameters across the four viruses, as well as PSRF estimates for the shared parameter $\nu$. Figure S7 illustrates the PSRF estimates for latent provincial and national at each time point. Both figures demonstrate that the MCMC algorithm has reached sufficient convergence, with most parameters having PSRF values below 1.02.

### S7 Comparison of latent concentration to clinical surveillance data

Figure S8 overlays latent provincial wastewater concentration with clinical test positivity for each province and virus (the national time-series is shown in the main manuscript), while Figure S9 and Figure S10 present the corresponding scatter plots with estimates of $R^2$ at the national and provincial level, respectively.

Table S5: Sites included in the window sensitivity analysis, each with a full two-year observation window.

| Province | Site | Site ID | Population | N obs. |
|---|---|---|---|---|
| British Columbia | Vancouver Annacis Island | VAI | 1419919 | 208 |
| British Columbia | Vancouver Iona Island | VII | 771638 | 205 |
| Manitoba | Winnipeg North End | WPN | 404000 | 517 |
| Manitoba | Winnipeg South End | WPS | 176000 | 512 |
| Ontario | Toronto Ashbridges Bay | TAB | 1447246 | 209 |
| Ontario | Toronto Humber | THU | 674800 | 209 |
| Quebec | Montreal North | MMN | 1018516 | 205 |
| Quebec | Montreal South | MMS | 1033404 | 207 |

### S8 Comparison of 1-year vs 2-year window sizes

In this analysis, three MCMC algorithms of the hierarchical model were conducted:

1. A data window from 1 January 2024 to 31 December 2025, reflecting the same 2-year window used in the main inference.

2. A data window from 1 January 2024 to 31 December 2024, a 1-year window representing the first half of the data used in the main inference.

3. A data window from 1 January 2025 to 31 December 2025, a 1-year window representing the second half of the data used in the main inference.

Sites with observed sample concentrations that span the entirety of the data window in the main inference were selected. Table S5 illustrates the sites used in this analysis. Figure S11 presents the results of this analysis. In this figure, estimates from the two 1-year windows are stitched together as a single time-series and compared to the estimates from the 2-year window.

Table S6: Site addition and removal events applied in the site-event sensitivity analysis.

| Province | Site ID | Site | Event | Date | % prov. pop. change |
|---|---|---|---|---|---|
| British Columbia | VII | Vancouver Iona Island | Removed | 1 January 2025 | -35.2 |
| Manitoba | WPS | Winnipeg South End | Added | 1 January 2025 | +43.6 |
| Ontario | THU | Toronto Humber | Removed | 1 October 2024 | -31.8 |
| Quebec | MMN | Montreal North | Added | 1 July 2024 | +98.6 |

### S9 Addition and removal of sites

The same subset of sites used in section S8 were used to test robustness of the hierarchical model. In this analysis, sites were "added" or "removed" by censoring observations in the dataset. Table S6 presents the site events that occurred in this analysis, and the subsequent change in the latent provincial population as a result of each event. Two MCMC algorithms were executed: one with no site events occurring; and another with the site events.

Figure S12 shows the magnitude of divergence varied between provinces and viruses; both the population share of the affected site and the concordance between its signal and that of the remaining provincial sites are potential factors explaining these differences.

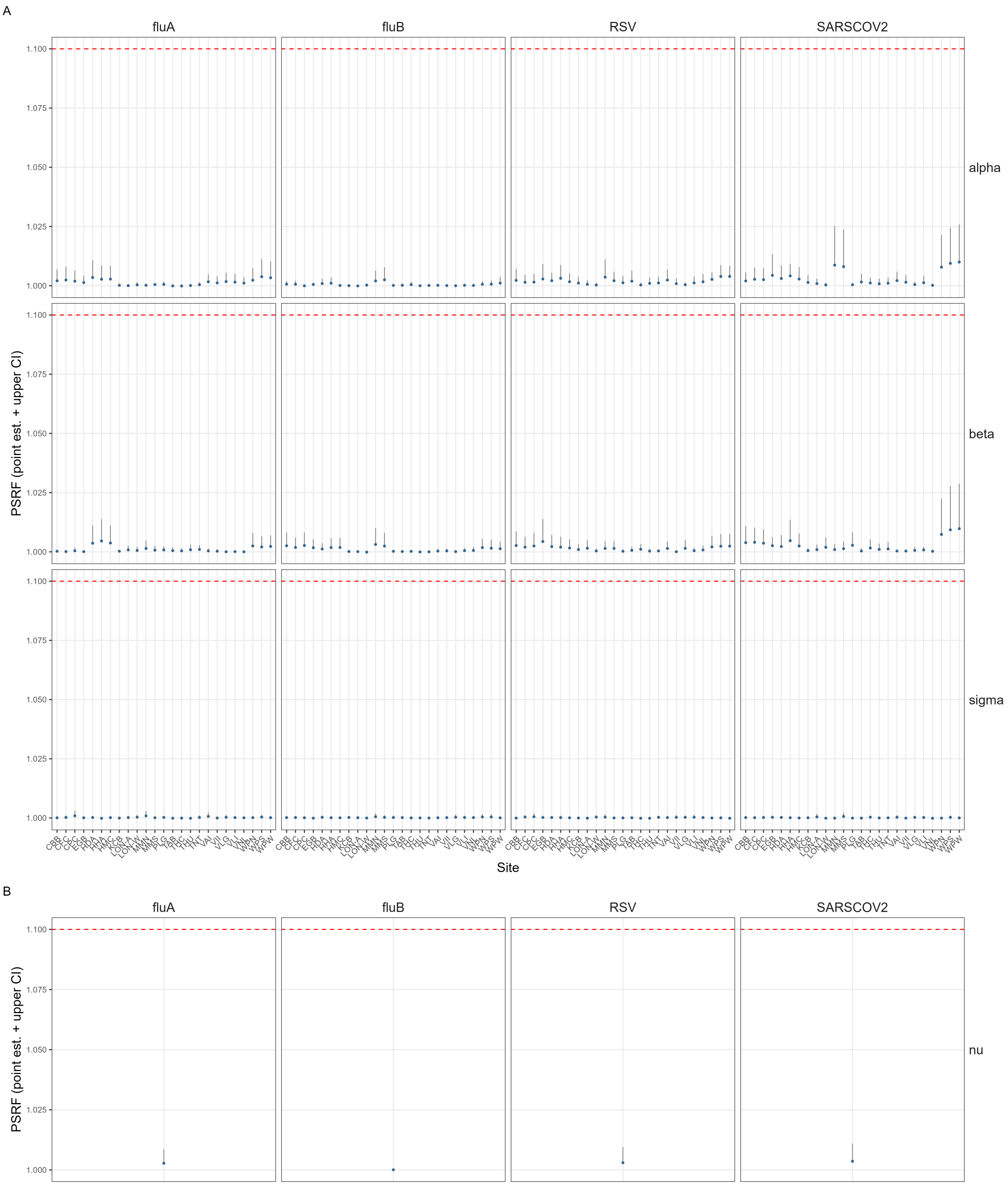


Figure S6: A) Estimates of potential scale reduction factor (PSRF) for site-level parameters $\alpha$, $\beta$, and $\sigma$ for SARS-CoV-2, Influenza A, Influenza B, and RSV across all sites used in the model. Each subplot represents site estimates (point estimate + upper bound) of PSRF for each virus and parameter. X-axis represents the site ID, while the y-axis represents the PSRF value. B) Estimates of PSRF for $\nu$ for SARS-CoV-2, Influenza A, Influenza B, and RSV.

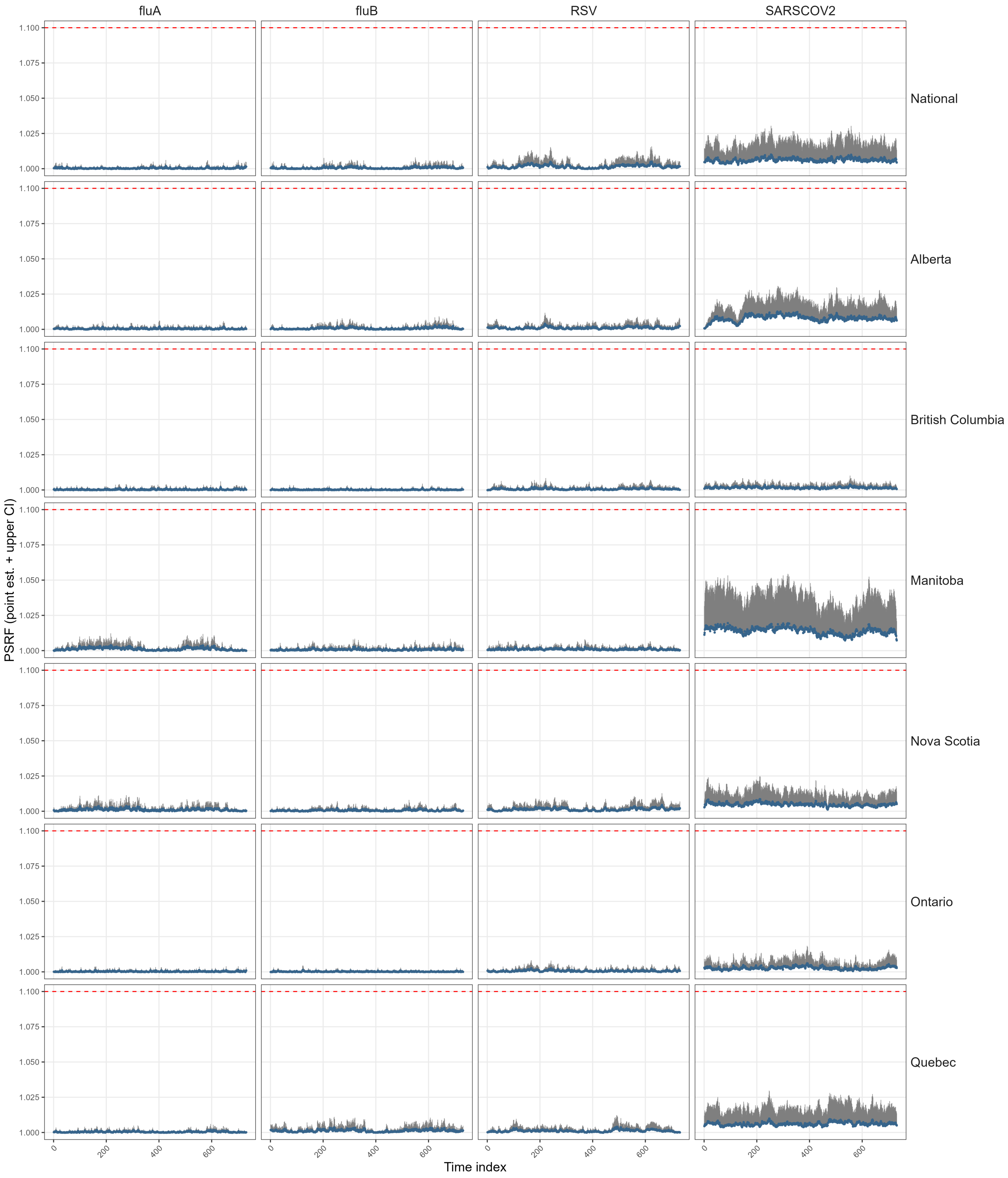


Figure S7: Estimates of potential scale reduction factor (PSRF) for national and provincial latent concentration each time point for SARS-CoV-2, Influenza A, Influenza B, and RSV. Each subplot represents PSRF estimates (point estimate + upper bound) for each location and virus. X-axis represents the time point, while the y-axis represents the PSRF value.

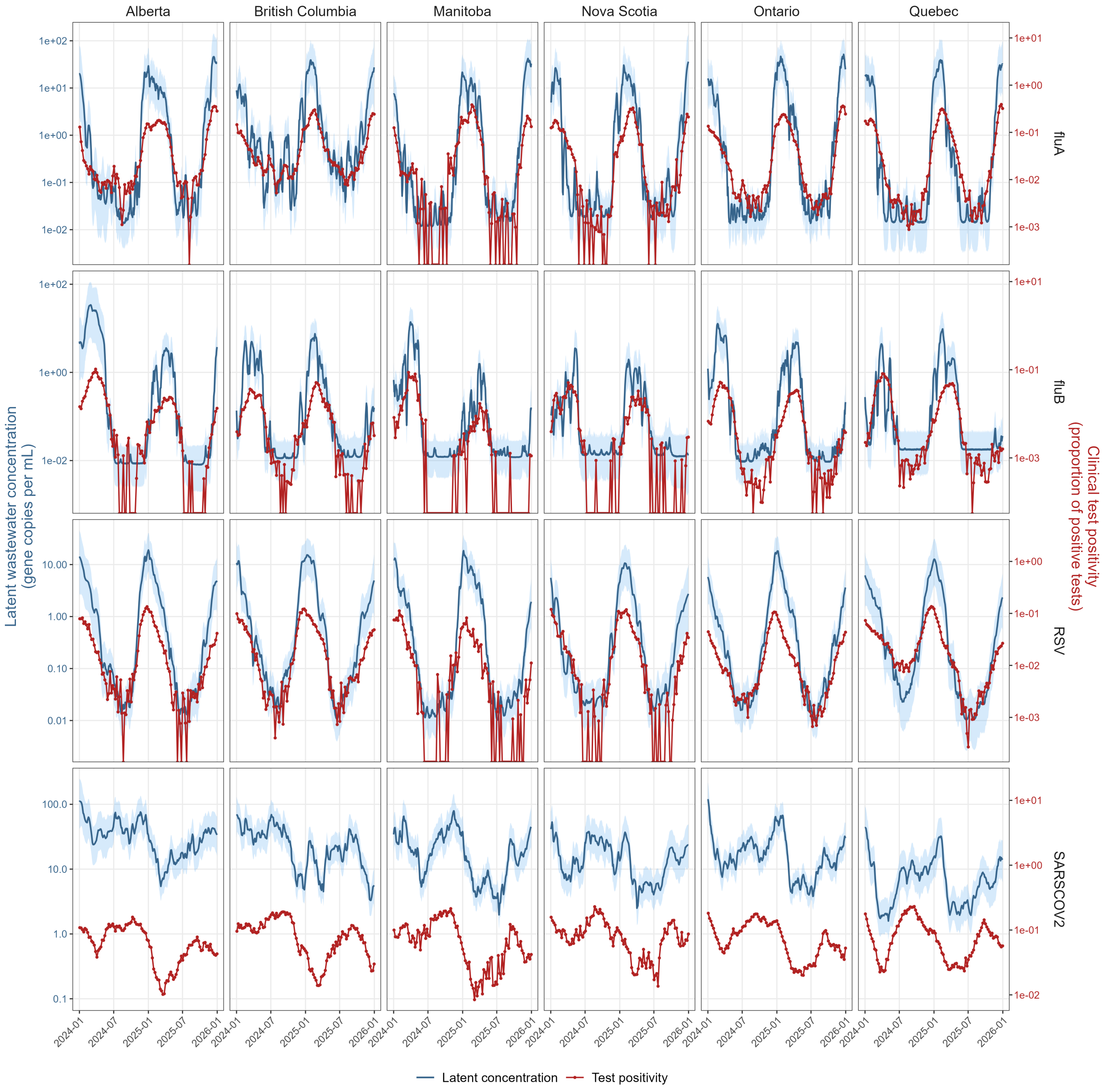


Figure S8: Comparison of latent provincial wastewater concentration and clinical test positivity for SARS-CoV-2, Influenza A, Influenza B, and RSV. Each subplot represents the time series for a given province and virus, with latent concentration (blue, left axis) shown as the posterior mean and 95% credible interval, and clinical test positivity (red, right axis) shown on matching weekly dates. The two series are displayed on separate axes to emphasize that wastewater concentration and test positivity are measured on different scales.

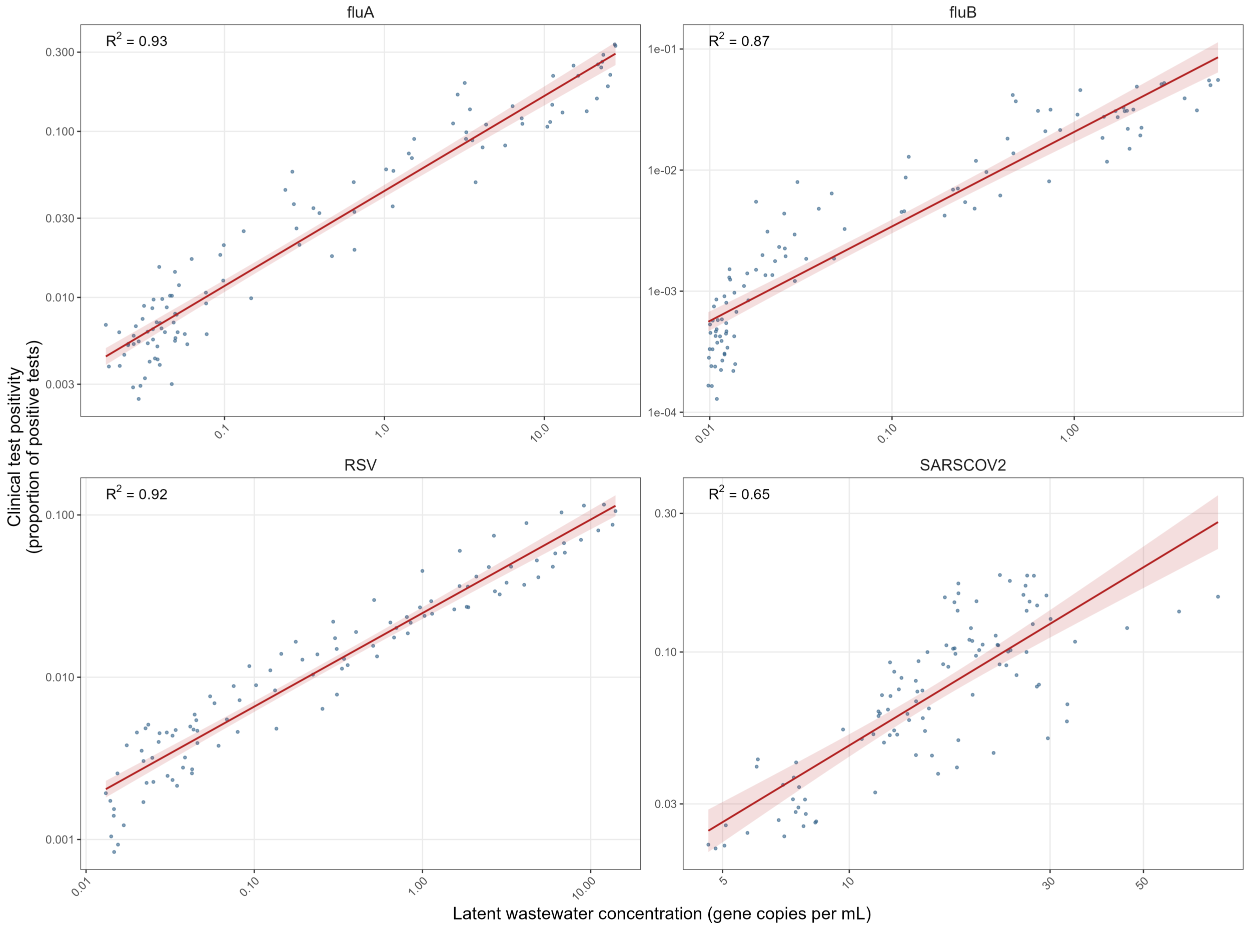


Figure S9: Association between latent national wastewater concentration and clinical test positivity for SARS-CoV-2, Influenza A, Influenza B, and RSV. Each subplot presents test positivity against mean daily latent concentration on matching dates, with a linear regression fit (red line and shaded 95% confidence interval). The coefficient of determination ($R^2$) from the regression of $\log_{10}$ test positivity on $\log_{10}$ latent concentration is annotated in each panel. The horizontal axis represents the latent concentration, while the vertical axis represents clinical test positivity, both axes on the log-scale.

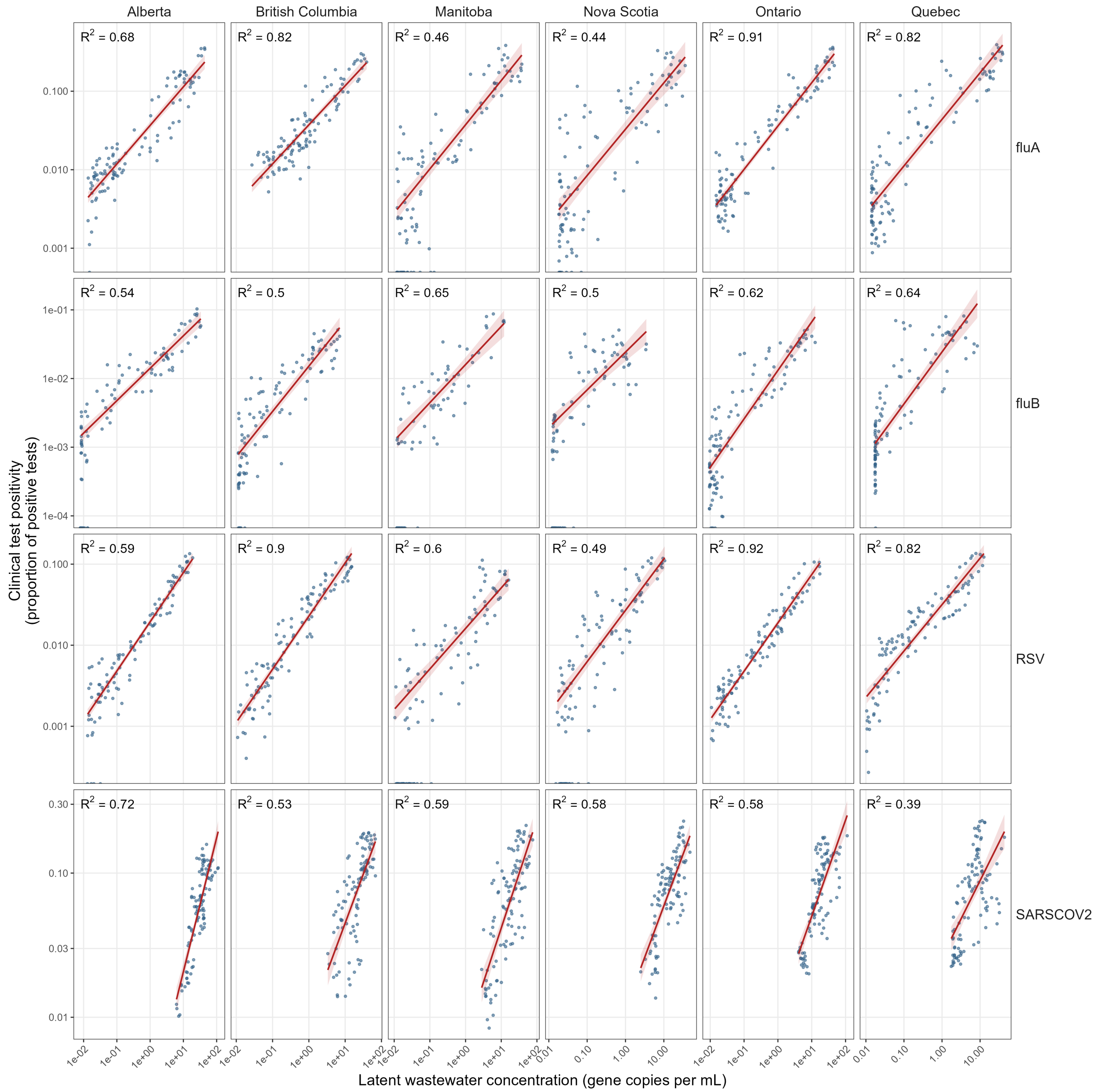


Figure S10: Association between latent provincial wastewater concentration and clinical test positivity for SARS-CoV-2, Influenza A, Influenza B, and RSV. Each subplot presents test positivity against mean daily latent concentration on matching dates for a given province and virus, with a linear regression fit (red line and shaded 95% confidence interval). The coefficient of determination ($R^2$) from the regression of $\log_{10}$ test positivity on $\log_{10}$ latent concentration is annotated in each panel. The horizontal axis represents the latent concentration, while the vertical axis represents clinical test positivity, both axes on the log-scale.

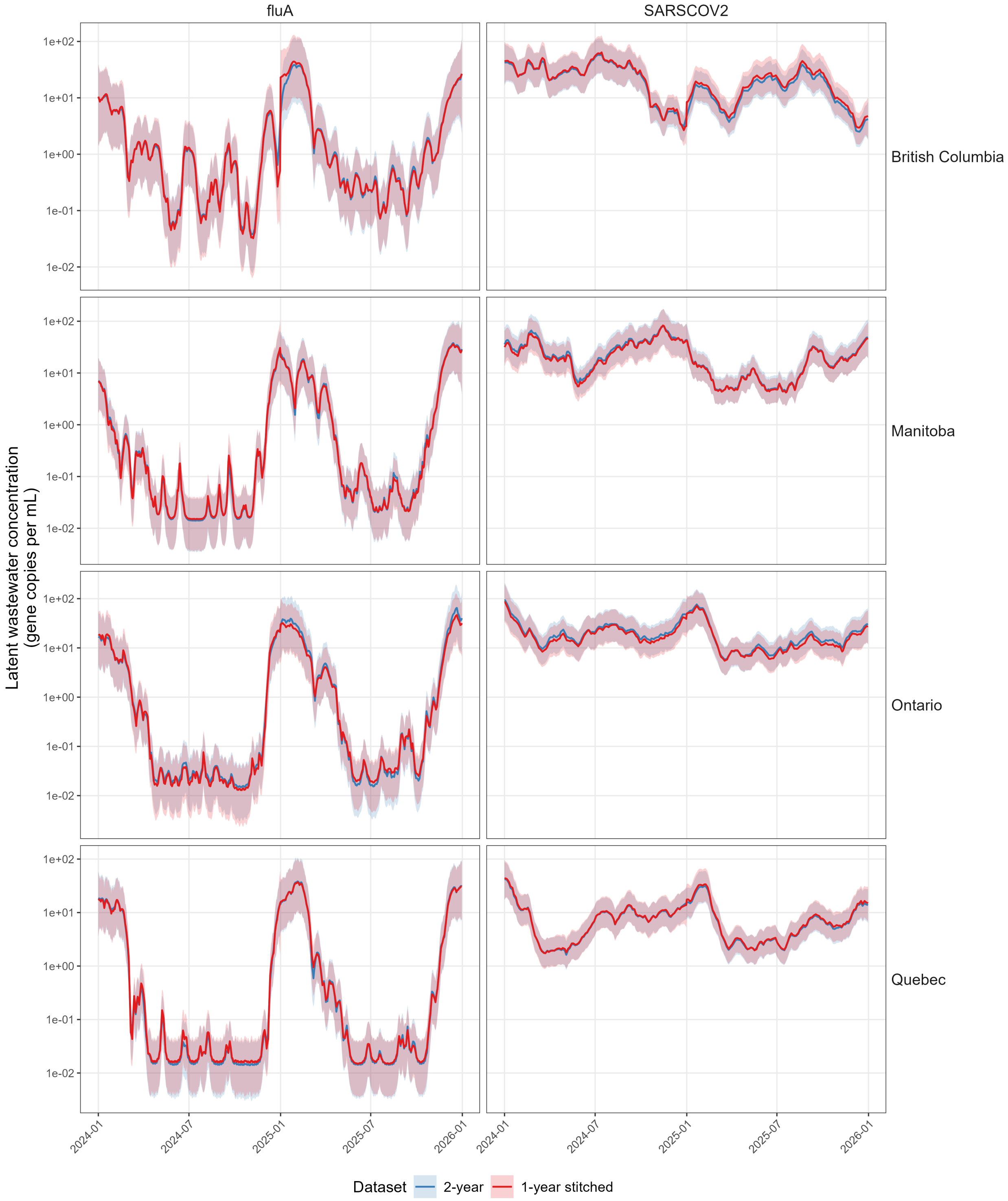


Figure S11: Window sensitivity analysis comparing provincial latent wastewater concentration estimated from the full 2-year data window (blue) against the estimate stitched together from two independent 1-year runs (red). Solid lines show the posterior mean and shaded ribbons the 95% credible interval, faceted by province (rows) and virus (columns).

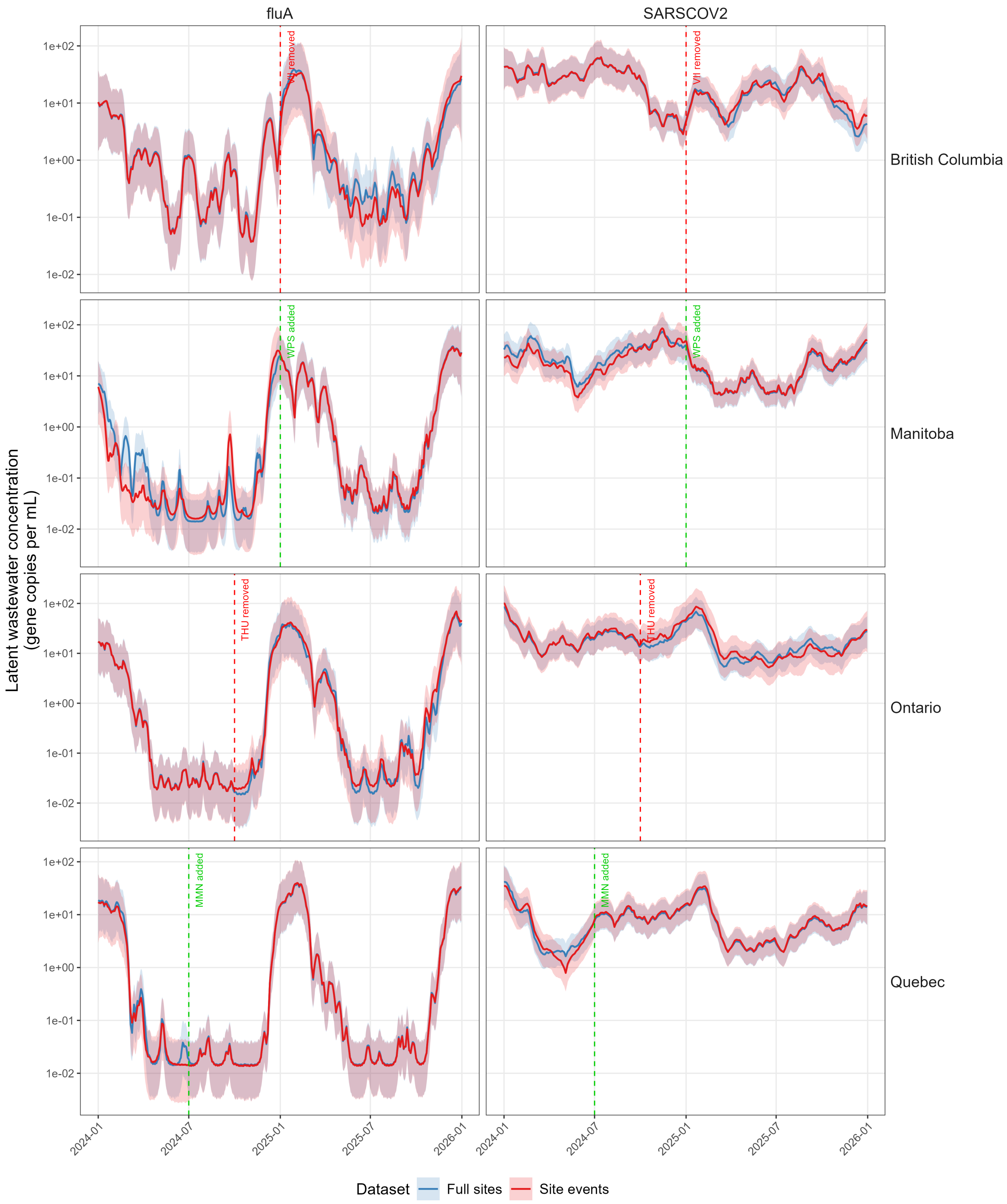


Figure S12: Site-event sensitivity analysis comparing provincial latent wastewater concentration estimated with all sites reporting throughout the window (blue) against an estimate where one site per province is added or removed partway through the window (red). Dashed vertical lines mark each site event (addition in green, removal in red) (Table S6). Solid lines show the posterior mean and shaded ribbons the 95% credible interval, faceted by province (rows) and virus (columns).
